# Effectiveness and cost effectiveness of a 12 month automated text message intervention for weight management in postpartum women with overweight or obesity: protocol for the Supporting MumS (SMS) multi-site, parallel-group, randomised controlled trial

**DOI:** 10.1101/2024.01.08.24300973

**Authors:** Dunla Gallagher, Eleni Spyreli, Annie S Anderson, Sally Bridges, Chris Cardwell, Elinor Coulman, Stephan U Dombrowski, Caroline Free, Suzie Heaney, Pat Hoddinott, Frank Kee, Cliona McDowell, Emma McIntosh, Jayne V Woodside, Michelle C McKinley

## Abstract

**Introduction:** The reproductive years can increase women’s weight-related risk. Evidence for effective postpartum weight management interventions is lacking and engaging women at this time is challenging. Following a promising pilot evaluation of the Supporting MumS intervention, we assess if theory-based and bidirectional text messages to support diet and physical activity behaviour change for weight loss and weight loss maintenance, are effective and cost-effective for weight change in postpartum women with overweight or obesity, compared with an active control arm receiving text messages on child health and development.

**Methods and analysis:** Two-arm, parallel group, assessor-blind randomised controlled trial with cost-effectiveness and process evaluations. Women (n=888) with body mass index ≥25 kg/m2 and within 24 months of giving birth, recruited via community and NHS pathways through five United Kingdom sites targeting areas of ethnic and socioeconomic diversity. Women are 1:1 randomised to the intervention or active control groups, each receiving automated text messages for 12 months. Data is collected at 0, 6, 12 and 24 months. The primary outcome is weight change at 12 months from baseline, compared between-groups. Secondary outcomes include weight change (24 months) and waist circumference (cm), proportional weight gain (>5kg), BMI (kg/m2), dietary intake, physical activity, infant feeding and mental health (12 and 24 months respectively). Economic evaluation examines health service usage and personal expenditure, health-related quality of life and capability wellbeing to assess cost-effectiveness over the trial and modelled lifetime. Cost-utility analysis examines cost per quality-adjusted life years gained over 24 months. Mixed method process evaluation explores participants’ experiences and contextual factors impacting outcomes and implementation. Stakeholder interviews examine scale-up and implementation.

**Ethics and dissemination:** Ethical approval obtained before data collection (West of Scotland Research Ethics Service REC 4 22/WS/0003). Results will be published via a range of outputs and audiences.

Trial Registration [2a]: ISRCTN16299220 (prospectively registered).

**Strengths and Limitations:**

- This is the first trial to examine the effectiveness and cost-effectiveness of a behavioural intervention for supporting weight loss in postpartum women with overweight or obesity that is delivered solely by text messages.
- The trial tests a novel evidence and theory-based text message intervention with embedded behaviour change techniques that is fully automated and incorporates two-way messaging to encourage engagement and delivery of specific behaviour change techniques.
- Researchers conducting participant recruitment and outcome data collection are blind to group allocation. Any situations where blinding is not maintained are recorded and reported. Due to the nature of the intervention, it is not possible to blind participants to randomisation.
- The trial includes an active control comparator to minimise disappointment bias and attrition related to randomisation experienced in previous weight loss intervention trials.(1)
- Home visits by the research team are offered to participants for data collection to facilitate participation in research assessments, helping to overcome barriers to taking part and address issues in participant retention seen in other postpartum trials.

**Administrative information:** This protocol is written in line with the SPIRIT checklist,(2) with recommended content indicated by the numbers in square brackets after section titles.

Title [1]: Effectiveness and cost effectiveness of a 12 month automated text message intervention for weight management in postpartum women with overweight or obesity: protocol for the Supporting MumS (SMS) multi-site, parallel-group, randomised controlled trial.

Protocol version [3]: Version 3.0 dated 10^th^ May 2023.

Trial status: Opened to recruitment in April 2022, with the first participant randomised in May 2022 and participant recruitment completed in May 2023. The trial is in follow-up until May 2025, with primary outcome data collection to be completed in May 2024.

## Introduction

### Background [6a]

This trial addresses the development of overweight and obesity across the childbearing years. Entering pregnancy with a high body mass index (BMI) increases many health risks for these mothers and their babies, including gestational diabetes, hypertensive disorders, caesarean section and birth complications, compared with recommended weight women.(3, 4, 5, 6) Excessive gestational weight gain (GWG) and postpartum weight retention (PPWR) are common (7, 8) and many women are at risk of further weight gain across the extended postpartum period.(9) By 18 months postpartum, about one in five women have moved into a higher BMI category (8) which increases the risk of complications in subsequent pregnancies and contributes to long-term overweight and obesity.(9)

Effective and appropriate weight management interventions in women during the postpartum period, which account for the well-recognised barriers for new mums such as time constraints and childcare, are lacking.(9, 10, 11, 12, 13, 14) Previous intervention studies are characterised by poor recruitment and high rates of attrition (1, 15, 16, 17, 18) and have not adequately considered the difficulties in reaching this population and specific barriers to lifestyle behaviour change that come with having a baby.(19, 20, 21, 22, 23) In-person and structured weight management approaches, such as those delivered in community and group settings, are unlikely to be successful with this population due to the commitment required, and such approaches may exacerbate health inequalities.(24, 25, 26, 27) More appropriate ways of recruiting and engaging with postpartum women to achieve sustained behaviour change are required (1, 9) and a need to evidence cost-effectiveness to inform implementation.(12)

Recent evidence, mostly from feasibility or pilot studies, indicates that technology usage shows promise in postpartum weight management, perhaps because this form of delivery fits more seamlessly with women’s lives at this stage.(17, 28, 29, 30) Mobile technologies can offer a more flexible and individualised ‘any time, any place’ approach to behavioural weight management interventions.(9) Text message interventions have high reach potential and allow for flexible scheduling and interactivity,(31) potentially making them convenient for mums with limited time and helpful for overcoming health inequalities. Systematic reviews support the use of text messaging in behavioural weight management interventions but indicate further evaluation is required.(32, 33, 34, 35, 36). They are cost-effective and readily scalable for changing other health behaviours.(37, 38)

The Supporting MumS (SMS) intervention, consisting of a library of text messages, was developed with personal and public involvement (PPI) to support weight management in the postpartum period.(28) Messages focus on diet and physical activity with embedded behaviour change techniques (BCTs) informed by behaviour change theory and evidence. In a previous pilot evaluation, one hundred women with overweight or obesity from Northern Ireland (NI), who had a baby within the previous 24 months, were recruited and randomised to receive text messages about weight loss (intervention group) or child health and development (active control) for 12 months. The study demonstrated the feasibility and acceptability of the intervention and an independent Trial Steering Committee (TSC) judged that the pre-specified progression criteria to proceed to a full randomised controlled trial (RCT) (i.e., successful recruitment, high retention and no differential attrition, high acceptability of the intervention, and evidence of positive indicative effects) were met.(28)

### Rationale [6a]

This RCT addresses the following research question: Is an automated 12 month text message intervention, designed to support weight loss and weight loss maintenance for postpartum women with overweight or obesity, effective and cost-effective for weight loss at 12 months, compared with an active control group receiving text messages on child health and development?

SMS is a parallel, 2-arm multi-site RCT using automated and bidirectional text messages to support weight management over a 12 month period in postpartum women with overweight or obesity who have had a baby in the last 24 months, comparing weight change at 12 months from baseline between the intervention and active control groups. This RCT protocol is based on the pilot RCT methodology, but changes were to remove: 1) use of a sealed pedometer to assess physical activity as this was not acceptable to women and, 2) a discussion forum intervention component as it was not used by pilot RCT participants. We added considerations for data collection in the event of COVID-19 restrictions. In moving from a single site pilot study to a multi-site RCT recruiting an ethnically and socioeconomically representative sample, we conducted a six-month period of intervention and recruitment method adaptation, with PPI, ensuring the broad acceptability and cultural relevance of the messages and enhancing the generalisability of the findings to different groups of women across the United Kingdom (UK), thus informing implementation.

The increased prevalence of obesity, its associated complications and weight management services, is costly to the National Health Service (NHS). NHS costs associated with caring for a pregnant woman with a BMI of ≥25 kg/m^2^ are up to 37% higher than caring for a woman with a healthy BMI.(39) Both National Institute for Health and Care Excellence (NICE) guidance (13) and an associated review of the cost-effectiveness of weight management interventions following childbirth,(40) outline the potential of postpartum interventions to be a cost-effective way of reducing the long-term risks of obesity, heart disease, cancer and diabetes, and recommend health professionals should advise women within two years of having a baby to eat a nutritious diet and keep active to encourage postpartum weight reduction.(13, 41)

Postpartum weight management interventions combining dietary and activity behaviour change have moderate positive influence on maternal weight (12, 15) and those including self-regulatory BCTs are likely to be more successful.(10, 12) The postpartum period could be an ideal time to intervene to shape new health behaviours as women may be motivated and receptive to health information.(42) However, women highlight a need for additional weight management support during the postpartum period as little is currently provided.(43)

The present trial allows women to take part up to 24 months postpartum. Qualitative evidence suggests that every woman’s postpartum journey is different (19, 20, 21, 22, 23) and the optimal time to engage women in postpartum weight management is currently unknown.(9, 11) Allowing this extended inclusion window accommodates different maternal weight trajectories across this time.(44, 45) Furthermore, women may prepare for another pregnancy during this period, so there is potential to support better health for subsequent pregnancies.(9)

Mobile phone usage and ownership are widespread (95% of UK households) amongst all sectors of society, irrespective of socioeconomic status.(31) Text messaging is a simple communication mode that does not require smartphone or tablet technology, unlike web-based interventions or apps. This supports health equity through high reach potential at a low cost, making it conducive with scale-up and implementation, unlike some weight loss interventions which can be resource intensive and expensive (46) and can exacerbate health inequalities.(47, 48) A text messaging intervention can be delivered flexibly and on a sustained basis, can be reactive and proactive, and does not necessarily rely on participant initiation.(31, 49) Using the real-time advantages offered by mobile technologies to deliver behavioural weight management support to postpartum women has the potential to encourage behaviours that may improve maternal health in both the short and long-term.(50) Trial results are relevant for other population groups given the possible advantages of this weight management approach. The NHS aims to embrace the potential of digital strategies for improving UK health (51) and text message interventions could complement any weight management support currently delivered across the NHS, to help ease the current burden on an over-stretched service.

The SMS trial adopts a fully automated approach to intervention delivery whilst still offering feedback to participants. The trial makes a novel and important contribution to the field of behavioural text message interventions in a number of ways: there are currently few such interventions that are fully automated, where text messaging is the main mode of delivery and two-way messaging is used to encourage engagement and delivery of specific BCTs; there are also few which consider both weight loss and weight loss maintenance, last 12 months or more and use an active control to minimise disappointment bias.(28)

The National Institute of Health and Care Research (NIHR) awarded further funding to explore postnatal mental health outcomes and needs to inform application of the priorities in the NHS Long Term Plan via existing studies. We added data collection methods related to mental health (postnatal depression, depression and anxiety, mother and child relationships, service and treatment experiences) in February 2022, allowing an opportunity to examine poor postnatal mental health prevalence rates, trajectories, predictors, inequalities and service needs, in an ethnically and socioeconomically diverse sample from all four UK countries. The trial also allows further validity and reliability testing of the Me and My Baby (MaMB)/Me and My Child (MaMC) screening tool which was co-produced by health visitors and the community to assess the mother and child relationship. Funding for this workstream was contracted in March 2022.

### Explanation for the choice of comparators [6b]

An active control comparator is used as pilot study findings indicate that this design aspect supports good participant retention, acceptability and satisfaction, where previous weight loss trials with postpartum women have reported high levels of attrition and disappointment bias in those allocated to no treatment or usual care control groups.(1) Active control participants are assessed for the same outcome measures as the intervention group at 6, 12 and 24 months. We advertise the purpose of the trial as testing two new text message services to minimise women’s preference for one group over another. The participant information sheet (PIS) clearly describes that women have a 50:50 chance of being assigned to receive messages about child health and development or messages about weight management, thus preserving participant autonomy.

### Objectives [7]

#### Primary objective

To conduct a 2-arm parallel-group RCT comparing weight change at 12 months for postpartum women with overweight or obesity who receive text messages about weight management with an active control.

#### Secondary objectives

- To assess differences between-groups in secondary outcomes.
- To assess the cost-effectiveness of the SMS intervention compared with an active control comparator.
- To assess the prevalence and trajectories of postnatal mental health in women across the UK, particularly those from marginalised groups, including assessment of the mother-child relationship.
- To conduct a process evaluation to explore women’s experiences of the intervention, the pathways through which the intervention effects are mediated and contextual factors affecting the outcomes or future implementation of the intervention.
- To seek permission for routine data linkage for long term health outcomes (mother and youngest child).
- To conduct a follow-up of women at 24 months (12 months after the intervention has ceased) to examine the long-term effect of the intervention and to conduct interviews with stakeholders to explore scale-up and implementation; this stage is contingent upon TSC and NIHR assessment of the 12 month primary outcome data and any other core outcome data required to make a fully informed decision about effectiveness and the clinical significance of weight change.

### Trial Design [8]

A pragmatic, multi-site, parallel, 2-arm, assessor-blind, 1:1 superiority RCT comparing weight change at 12 months for the SMS intervention and active control groups, with mixed methods process evaluation, cost-effectiveness modelling, consent for future data linkage to longer-term health outcomes for mothers and babies, 24 month participant follow-up (i.e. 12 months post-intervention) and implementation consultations with stakeholders.

## Methods and analysis

### Trial setting [9]

A multi-site trial with five recruitment sites in the UK: 1) Belfast, Northern Ireland (trial co-ordinating site); 2) Stirling, Scotland; 3) Cardiff, Wales; 4) London, England; and 5) Bradford, England. Recruitment sites were selected to target geographical areas of ethnic and socioeconomic diversity for the purposes of participant recruitment via community and NHS pathways.

### Eligibility criteria [10]

Women (in accordance with the NICE Postnatal care guideline NG1941, the term ’woman’ used in this trial includes people who do not identify as women but who are pregnant or have given birth) are eligible to participate in the trial if they are aged 18 years or over, have a BMI of ≥25 kg/m^2^, have had a baby within the last 24 months and own a mobile phone to allow them to receive personal text messages.

Women are excluded if their baby is less than 6 weeks old or if they have insufficient English language to understand short written messages, if they have had or plan to have any type of weight loss surgery, if they report having ever received a diagnosis of anorexia nervosa or bulimia from a doctor, or, if they are on a specialist diet and receiving dietetic care. Women are excluded if they are participating in any other weight management research study/programme currently or in the previous three months. Women are not eligible if they are pregnant and any consented participant becoming pregnant during the trial is excluded, as the intervention has not been designed for use in pregnancy.

### Interventions

#### Supporting MumS (SMS) intervention [11a]

A full description of the SMS intervention development has been reported elsewhere.(28) The intervention is described here in accordance with the Template for Intervention Description and Replication (TIDieR) guidance.(52) The intervention group receive fully automated text messages, including bidirectional and interactive features, about weight loss and maintenance of weight loss for 12 months. The evidence and theory-based intervention consists of a library of text messages focused on diet and physical activity with embedded BCTs known to be positively associated with weight management. The intervention logic model and sample intervention and active control text messages are available in supplemental appendix 1. Examples of how BCTs were incorporated into the messages is available elsewhere.(28)

##### Why-theory and components

The intervention is based on the Health Action Process Approach (HAPA) and a systematic review of over 100 behavioural theories which synthesised theoretical explanations for maintenance of behaviour change.(53) The intervention encourages a self-guided approach to weight-related behaviour change, as supported by the literature.(54, 55, 56, 57) ^T^he intervention focuses on dietary intake and physical activity to address energy balance related behaviour, with embedded evidence-based BCTs specifically linked to the relevant phases and psychological processes of behaviour change alongside consideration of barriers for this population.(28)

The messages adopt a friendly tone including humour to foster engagement and provide information, practical tips and advice including quotes from other mothers. They signpost to external resources and provide encouragement and motivation, discourage guilt and promote self-reflection. A core library of text messages (n= 353) to be delivered by the automated system was created. These include a weekly text message asking women to self-weigh and report their weight (n=50) and bidirectional messages to encourage engagement, self-monitoring, relapse prevention and to provide feedback, as follows: 1) “Yes/No” questions (n=36) which trigger an automated response to the participant based on their reply, 2) trigger words (‘slip-up’; ‘crave’; ‘bad day’ or ‘tired’) which can be texted by women at any time to prompt a reply designed to address these barriers and prevent relapses, 3) During months 7-12 when the focus shifts to maintenance strategies, the weekly text message asking women to report their weight asks them to add the word ‘up’ or ‘down’ or ‘same’ in relation to how their weight compares with the week prior. These keywords trigger an automated response. Responses for bidirectional messages are sent from pre-loaded message banks and are not individually scripted or tailored.

Women can also opt-in to receive messages related to weight management when breastfeeding (n=10) or stopping smoking (n=15). These messages are intended to alleviate participant concerns in relation to weight management whilst breastfeeding and to address vulnerabilities around weight gain during smoking cessation.

##### What-materials and procedures

The intervention consists of 353 core messages delivered to all women allocated to this group, in addition to messages associated with the interactive and optional components described above.

Social support is facilitated throughout the intervention using a “buddy system” (informed by a successful smoking cessation text message intervention (38)) where a participant can nominate a friend or family member to receive the same messages they are receiving, putting that person in a better position to offer support. Participants are sent instructions and reminders on how to do this within the core messages, along with information on the value that support can play in weight loss success. A ‘buddy’ can be requested at any stage throughout the intervention period.

##### Who-intervention provider

Intervention costs are provided by the NI Public Health Agency (PHA). Intervention delivery is fully automated via the existing pilot study text message platform developed by the London School of Hygiene and Tropical Medicine (LSHTM). The intervention package is designed so that it can be readily delivered by other text message providers to facilitate future implementation by the NHS or public services.

##### How - mode of delivery

Message exchange with participants is managed via a secure server at the LSHTM, using Firetext (58) to distribute messages to participants.

##### Where– setting

The intervention can occur any place and any time. Participants require a mobile phone with the function to receive text messages.

##### When and how much

Text message frequency starts at 14-15 sent per week during months one and two, tapering to 9-10 per week in months three to six. Fewer messages, four to five per week, are sent in the weight loss maintenance phase (months 7 to 12) when the emphasis shifts to reinforcing self-regulation techniques developed during the first six months of the intervention and encouraging maintenance relevant strategies such as relapse prevention.

The system delivers messages any time between 10am and 11pm, with messages programmed to arrive at varied times to avoid predictability. Participants can choose to block an additional period of the day that they want to be kept message-free e.g., 10am to 5pm.

Following randomisation, women allocated to the intervention group are programmed to start receiving messages on the subsequent Monday which schedules the weekly message asking participants to report their weight to be sent on Fridays.

### Active control [11a]

The active control messages were also developed by the trial team with PPI input (28), for mothers with babies aged six weeks up to 36 months (based on participant inclusion criterion). The messages relate to general childcare and development, with content consistent with evidence-based information provided by the NHS Start4life service (59) including weekly play ideas/activities and information on milestones, home safety, separation anxiety and similar topics. Active control messages do not address the intervention target or active ingredients (i.e. diet, physical activity and BCT content related to weight loss) nor do they prompt bidirectional functionality. Women receive three messages each week over 12 months with messages corresponding to the age of their baby, i.e., if their baby is six months old at randomisation, the participant starts to receive messages corresponding to this age.

Women allocated to the control group are offered to receive a booklet summarising the intervention content on completion of the trial.

### Criteria for discontinuing or modifying allocated interventions [11b]

Participants in both groups have the ability to pause, stop and restart the messages throughout by sending an instruction to the text system e.g., ‘STOP’, ‘START’. Researchers ask women choosing to discontinue the messages to provide any reason for doing so, to inform the process evaluation. Participants discontinuing messages remain in the trial unless they formally withdraw from the trial assessments.

### Strategies to improve adherence to interventions [11c]

Text message delivery is fully automated and centrally delivered via the LSHTM system which is intended to ensure treatment fidelity. Regular monitoring of the text message system is completed by the system developer and trial team members (CI and clerical officer) to check the content of sent and received messages (e.g., ensure web links are working, review spontaneous participant responses). Intervention engagement is assessed via text messages received from participants in response to the weekly weight prompt and the replies to bidirectional messages as described in [11a], self-report items in questionnaires at 6, 12 and 24 months and qualitative interviews at 6 and 12 months.

### Relevant concomitant care permitted or prohibited during the trial [11d]

Women who become pregnant during the trial will be excluded, as text message content has not been designed for pregnancy and primary outcome data (weight) can only be collected from women who are not pregnant. This is a self-directed intervention and women are permitted to access any other weight-loss services via NHS, voluntary or private sector pathways. Details of engagement with other services are captured at each data collection timepoint. Any weight loss surgery undergone by participants during the trial, and prescribed or over-the-counter medications that may affect weight, are documented at 6, 12 and 24 months to assess the impact on trial outcomes.

### Outcomes [12]

Trial outcomes, measures and the assessment schedule at 0, 6, 12 and 24 months are outlined in Table 2.

➢ *Primary outcome:* between-group difference in mean weight change (kg) from baseline to 12 months.
➢ *Secondary outcomes:* between-groups difference in mean weight change (kg) from baseline to 24 months and between-groups differences in mean waist circumference (cm), mean BMI (kg/m^2^), in the proportions of women gaining a substantial amount of weight (>5kg), dietary intake (Fat and Fibre Barometer plus questions on sugar intake), physical activity (IPAQ-SF) and infant feeding practices (Infant Feeding Survey), by the 12 and 24 month follow-up timepoints respectively.
➢ *Health economic outcomes:* NHS resource use and personal expenditure (GP usage, hospitalisations, prescribed and over-the-counter medications, lifestyle products and services, expenditure on food and drink, alcohol and smoking products); health-related quality of life (EQ-5D-5L with visual analogue scale) and Capability (ICECAP-A).
➢ *Process Evaluation:* to explore participants’ experiences and interactions with the intervention by site, ethnicity and socioeconomic status, including evaluating dose (number of women who choose to receive all text messages and those who pause or stop the messages), engagement with the intervention (text message responses and qualitative data), reach (trial records of recruitment, loss to follow-up and withdrawal) and acceptability (participants’ satisfaction ratings and qualitative data).
➢ *Exploratory outcomes:* moderator and mediator analysis (Table 2)
➢ *Qualitative sub-study:* telephone interviews with a purposive sample of participants at 6 and 12 months to understand the experiences of women receiving the intervention and active control text messages and the contextual factors related to engagement including postnatal mental health, to inform trial and process evaluation outcomes.
➢ *Stakeholder consultation:* if the intervention is shown to be effective at 12 months, stakeholder interviews will be conducted to explore factors relevant to intervention scalability and implementation, including any further work needed to develop a coherent implementation model.

**Table 2:**
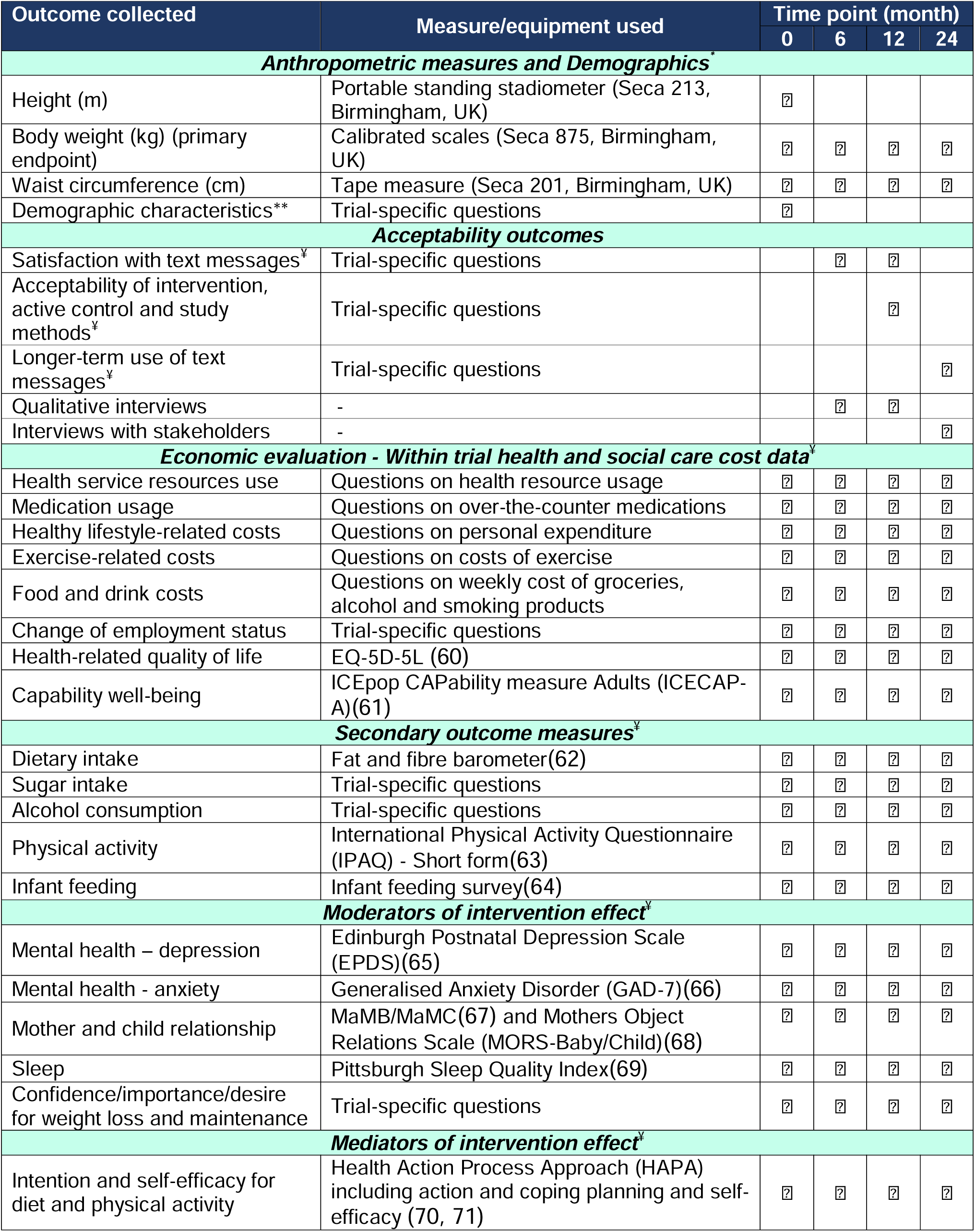
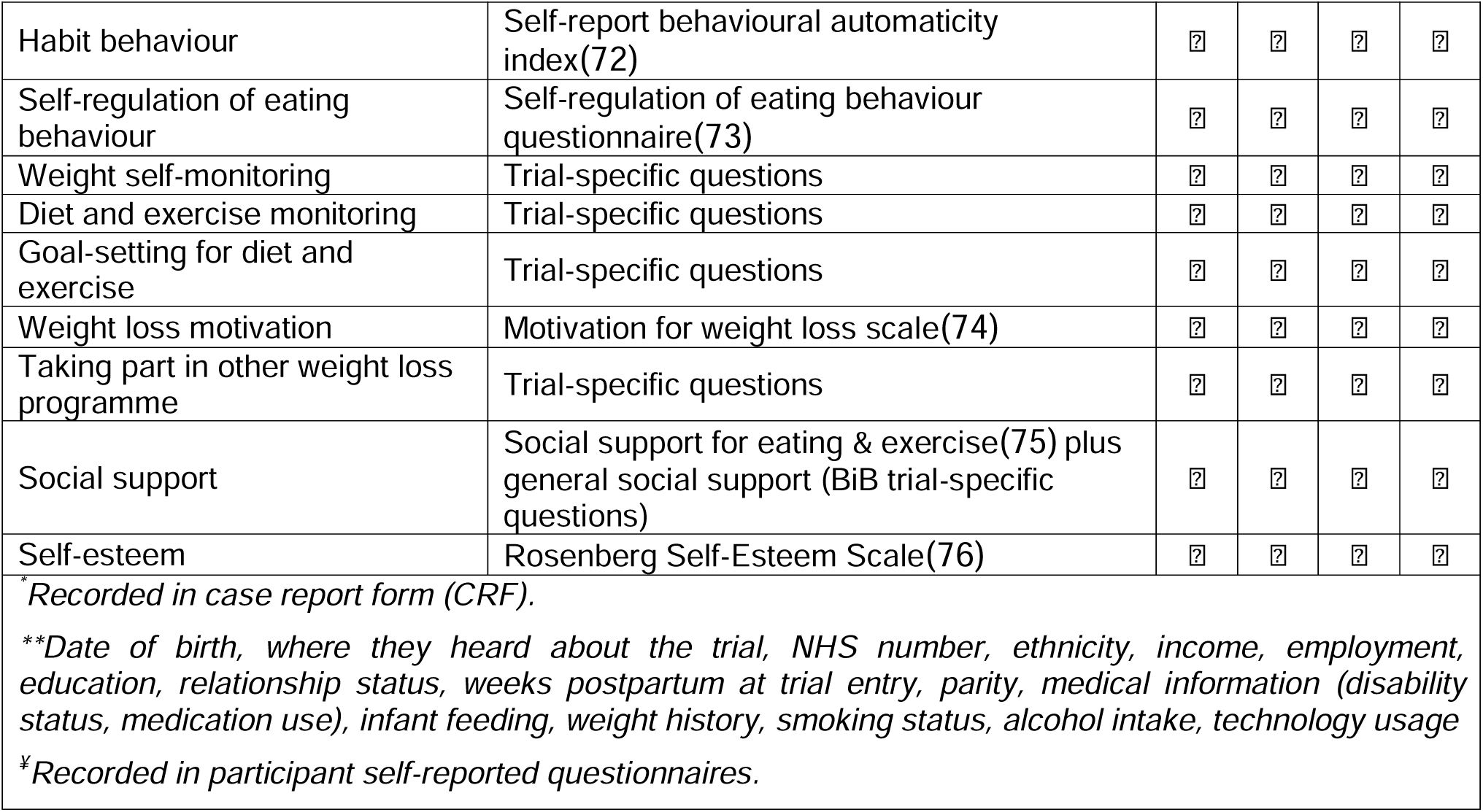
Supporting MumS trial assessments.

| Outcome collected | Measure/equipment used | Time point (month) |  |  |  |
| --- | --- | --- | --- | --- | --- |
|  |  | 0 | 6 | 12 | 24 |
| Anthropometric measures and Demographics* |  |  |  |  |  |
| Height (m) | Portable standing stadiometer (Seca 213, Birmingham, UK) | ? |  |  |  |
| Body weight (kg) (primary endpoint) | Calibrated scales (Seca 875, Birmingham, UK) | ? | ? | ? | ? |
| Waist circumference (cm) | Tape measure (Seca 201, Birmingham, UK) | ? | ? | ? | ? |
| Demographic characteristics** | Trial-specific questions | ? |  |  |  |
| Acceptability outcomes |  |  |  |  |  |
| Satisfaction with text messages‡ | Trial-specific questions |  | ? | ? |  |
| Acceptability of intervention, active control and study methods‡ | Trial-specific questions |  |  | ? |  |
| Longer-term use of text messages‡ | Trial-specific questions |  |  |  | ? |
| Qualitative interviews | - |  | ? | ? |  |
| Interviews with stakeholders | - |  |  |  | ? |
| Economic evaluation - Within trial health and social care cost data‡ |  |  |  |  |  |
| Health service resources use | Questions on health resource usage | ? | ? | ? | ? |
| Medication usage | Questions on over-the-counter medications | ? | ? | ? | ? |
| Healthy lifestyle-related costs | Questions on personal expenditure | ? | ? | ? | ? |
| Exercise-related costs | Questions on costs of exercise | ? | ? | ? | ? |
| Food and drink costs | Questions on weekly cost of groceries, alcohol and smoking products | ? | ? | ? | ? |
| Change of employment status | Trial-specific questions | ? | ? | ? | ? |
| Health-related quality of life | EQ-5D-5L (60) | ? | ? | ? | ? |
| Capability well-being | ICEpop CAPability measure Adults (ICECAP-A)(61) | ? | ? | ? | ? |
| Secondary outcome measures‡ |  |  |  |  |  |
| Dietary intake | Fat and fibre barometer(62) | ? | ? | ? | ? |
| Sugar intake | Trial-specific questions | ? | ? | ? | ? |
| Alcohol consumption | Trial-specific questions | ? | ? | ? | ? |
| Physical activity | International Physical Activity Questionnaire (IPAQ) - Short form(63) | ? | ? | ? | ? |
| Infant feeding | Infant feeding survey(64) | ? | ? | ? | ? |
| Moderators of intervention effect‡ |  |  |  |  |  |
| Mental health – depression | Edinburgh Postnatal Depression Scale (EPDS)(65) | ? | ? | ? | ? |
| Mental health - anxiety | Generalised Anxiety Disorder (GAD-7)(66) | ? | ? | ? | ? |
| Mother and child relationship | MaMB/MaMC(67) and Mothers Object Relations Scale (MORS-Baby/Child)(68) | ? | ? | ? | ? |
| Sleep | Pittsburgh Sleep Quality Index(69) | ? | ? | ? | ? |
| Confidence/importance/desire for weight loss and maintenance | Trial-specific questions | ? | ? | ? | ? |
| Mediators of intervention effect‡ |  |  |  |  |  |
| Intention and self-efficacy for diet and physical activity | Health Action Process Approach (HAPA) including action and coping planning and self-efficacy (70, 71) | ? | ? | ? | ? |
| Habit behaviour | Self-report behavioural automaticity index(72) | ? | ? | ? | ? |
| Self-regulation of eating behaviour | Self-regulation of eating behaviour questionnaire(73) | ? | ? | ? | ? |
| Weight self-monitoring | Trial-specific questions | ? | ? | ? | ? |
| Diet and exercise monitoring | Trial-specific questions | ? | ? | ? | ? |
| Goal-setting for diet and exercise | Trial-specific questions | ? | ? | ? | ? |
| Weight loss motivation | Motivation for weight loss scale(74) | ? | ? | ? | ? |
| Taking part in other weight loss programme | Trial-specific questions | ? | ? | ? | ? |
| Social support | Social support for eating & exercise(75) plus general social support (BiB trial-specific questions) | ? | ? | ? | ? |
| Self-esteem | Rosenberg Self-Esteem Scale(76) | ? | ? | ? | ? |
| <sup>*</sup>Recorded in case report form (CRF).<br><sup>**</sup>Date of birth, where they heard about the trial, NHS number, ethnicity, income, employment, education, relationship status, weeks postpartum at trial entry, parity, medical information (disability status, medication use), infant feeding, weight history, smoking status, alcohol intake, technology usage<br><sup>‡</sup>Recorded in participant self-reported questionnaires. |  |  |  |  |  |

### Participant timeline [13]

The trial flowchart includes the participant timeline for the RCT (Figure 1).

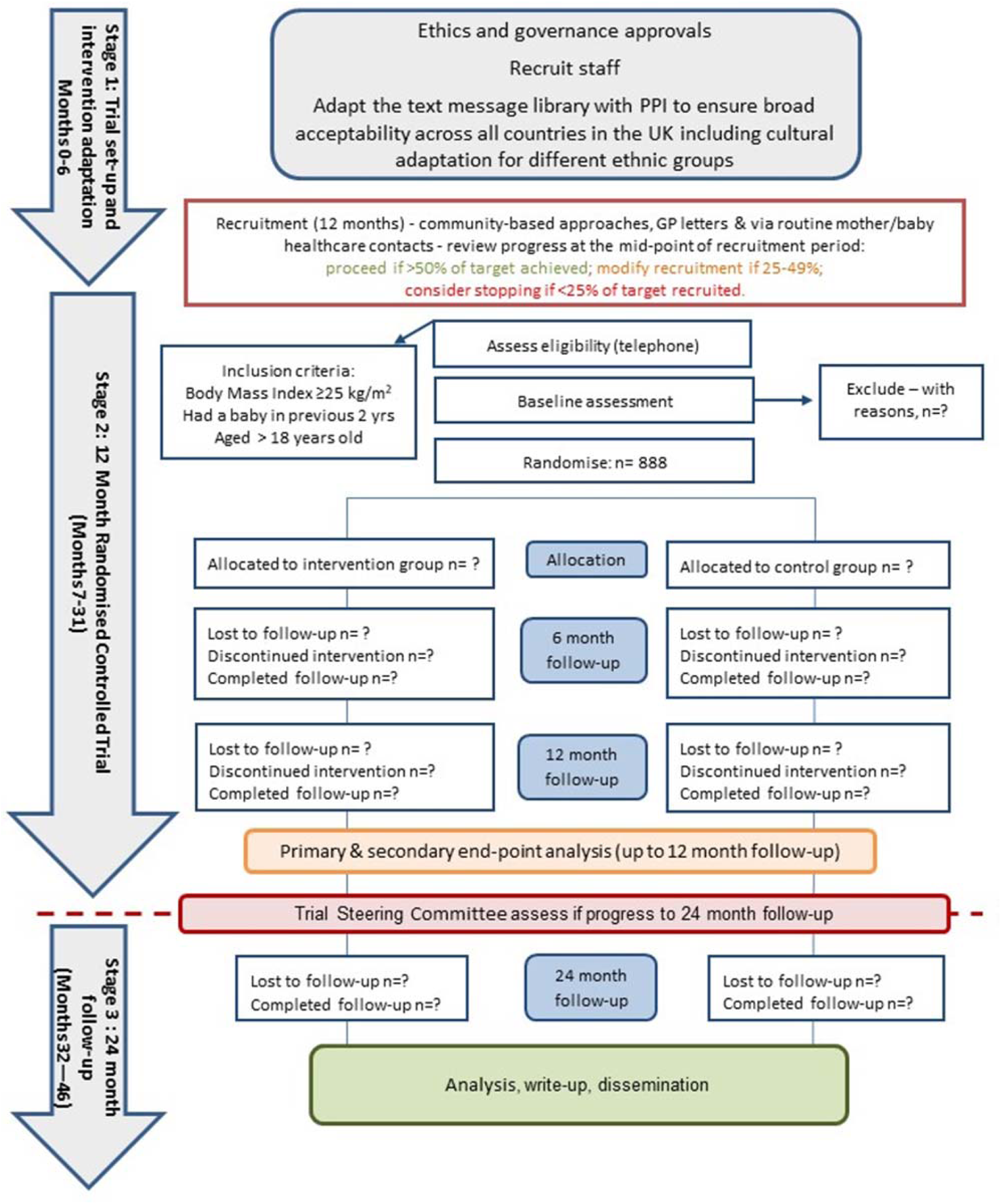

### Sample size [14]

The target sample size is 888 women over 12 months, with site-specific targets across the five geographical areas (London and Bradford, n=189, respectively; Belfast, Stirling and Cardiff, n=170, respectively). Pregnancy-related withdrawals are not replaced unless the loss to follow-up (12%) or pregnancy rates (15%) observed in the pilot RCT (28) are exceeded by the 6 month timepoint, which is monitored during Project Management Team (PMT) meetings.

#### Sample size calculation

The pilot trial showed a mean weight loss of 1.75kg in the intervention group compared with a mean gain of 0.19kg in the active control group between baseline and 12 months, equating to a between-groups difference in mean weight change, adjusted for baseline, of -1.67 kg (95% CI -4.88 to 1.55)).(28) Based on pilot data for the active control group and a standard deviation of 7.5kg, 594 completing participants (297 per group) are required to give the trial over 90% power to detect a statistically significant difference of 2kg in mean weight change from baseline, at the 5% level, between-groups. A mean difference of 2kg is accepted as being associated with metabolic health benefits and is often used to power weight loss studies.(57)

We observed a 15% pregnancy dropout rate in the pilot study, based on a 99% white ethnicity population.(28) Data from South Asian women in the Born in Bradford’s Better Start (BiBBS) cohort (77) indicates that pregnancy dropout rates could be higher in more ethnically diverse samples, around 22%. To have 119 participants completing per site, and accounting for a loss to follow-up rate of 15% and higher anticipated pregnancy exclusion rates in areas with greater ethnic diversity, the following site-specific sample sizes were calculated: 1) Belfast, Stirling and Cardiff; 15% pregnancy rate + 15% loss to follow-up = 170 women, 2) Bradford and London; 22% pregnancy rate + 15% loss to follow-up= 189 women. Therefore, the total sample size for the trial is 888 women (444 per group).

### Recruitment [15]

Based on successful pilot study recruitment processes,(28) trial information is disseminated via community settings likely to be attended by mothers of young children (e.g., parent and toddler groups, libraries, breastfeeding support groups), social media and through contacts with healthcare professionals and GP practices, including both routine appointments and contacting potentially eligible women identified using patient database searches. A range of recruitment pathways are used to target ethnically and socioeconomically diverse populations.

In Bradford, principal trial recruitment is through BiBBS, an existing birth cohort study, with trial information disseminated to participants who previously consented to be informed of other relevant research studies.

#### Monitoring recruitment progress

Based on the pilot study recruitment rate,(28) we plan to recruit the target sample over a 12 month period. We employ a recommended traffic light system (Figure 1) to review trial progression at the mid-point of the recruitment period,(78) with the following actions: proceed if >50% of target achieved; modify recruitment approach if 25-49% achieved and consider stopping (discuss with funder and TSC) if <25% of target recruited.

### Screening [10]

All trial information materials disseminated via described recruitment pathways, direct women who are potentially interested in taking part to contact the research team via telephone, email or by accessing the trial website/scanning a QR code to submit a secure pro-forma with their contact details.

Site researchers contact women expressing an interest in their area to provide a PIS, privacy notice and copy of the consent form (supplemental appendix 2) via email or post. Audio versions of the PIS and privacy notice are available on the trial website to increase accessibility. To comply with the Welsh Language Act 1993, Welsh language versions of the PIS and Consent Forms are available, on request. Women are given at least 48 hours to consider the information before being contacted again by telephone by the research team to discuss the trial procedures and any queries then invited to take part. Up to three contact attempts are made to invite all women expressing interest for screening. BMI eligibility is initially determined using women’s self-reported height and weight but later measured and verified by the researcher prior to collecting informed consent. Women who are deemed ineligible due to their baby being less than six weeks old will be invited for rescreening when they reach six weeks postpartum.

### Informed consent and withdrawal [26a]

For women meeting the eligibility criteria and willing to proceed with trial participation, a baseline researcher visit is arranged. Home visits are the primary approach to data collection, with women also offered the option of attending another venue if preferred (University building, community venue or their workplace).

Research staff, trained in Good Clinical Practice (GCP) guidelines, obtain written informed consent prior to collecting baseline data. At the baseline visit, women are offered the opportunity to ask further questions and confidentiality of data is explained. Informed consent is then taken. Women are reminded that they have the freedom to choose whether to take part or not. If willing to proceed, women are asked to initial and sign the trial consent form, with optional consent to be invited for telephone interview participation at 6 and 12 months. During the 6 and 12 month trial visits, women providing this optional consent at baseline are asked if they are still happy to be contacted to take part in a telephone interview. Participants completing telephone interviews are asked to provide their verbal consent for audio recording and for anonymised quotations to be used in trial publications before commencing interviews.

Once consented, participants are advised that they can withdraw from the trial at any time, without giving a reason, should they wish. Withdrawals are documented in the CRF along with any reason given for withdrawing. Participants excluded due to pregnancy are given the option to continue receiving the text messages. Anyone choosing this option needs to complete and return a pregnancy form to acknowledge that the messages have not been designed for pregnancy but may be useful after pregnancy. Data collected up to the point of withdrawal is retained for analysis unless the participant specifies otherwise.

### Additional consent provisions for collection and use of participant data and biological specimens [26b]

Optional aspects of trial consent include permission to be informed of future follow-up to this trial or other relevant research studies and permission for future data linkage to routinely collected health records for long term health outcomes (mother and youngest child) which is not within the current trial timeline. No biological specimens are collected.

### Assignment of interventions: allocation

#### Sequence generation [16a]

Participants are randomised in blocks of size 4 and randomisation is stratified by site, with the allocation sequence generated in STATA (using ralloc) by an independent statistician.

#### Concealment mechanism and allocation implementation [16b and 16c]

The allocation sequence is sent directly to the platform manager at LSHTM for upload to the secure web-based text message delivery system. After collecting consent and baseline data, site researchers enrol participants using the LSHTM randomisation system linked directly to the text message platform, implementing the random allocation sequence by accordingly sending automated intervention or control messages to the participant. Participants become aware of their group allocation when they start to receive the text messages; researchers do not have access to this information.

### Blinding [17a]

Researchers responsible for recruitment and collecting outcome data are blinded to group allocation. It is not possible to blind participants, but they are requested not to discuss the text messages they are receiving with researchers at each visit. This worked well in the pilot study with blinding maintained.(28) All statistical analyses are conducted blinded to group allocation until analyses of the 12 month data point are complete at which point the randomisation sequence is provided by the independent statistician who prepared it. We record any circumstances in which unblinding has occurred. The trial clerical officer, Chief Investigator and LSHTM text message system developer are not blind; these individuals see what group individuals are assigned to on the text message system for the purposes of monitoring the intervention delivery and the clerical officer is responsible for preparing the group-specific 12 month questionnaires. Blinded researchers do not see these questionnaires until after primary outcome data is collected. Interviews at 6 and 12 months are carried out by a researcher who has not had any previous contact with the participant.

Group allocation information for that participant is provided to the researcher before the interview so the correct interview guide can be used.

### Emergency unblinding [17b]

Given the nature of the intervention, emergency unblinding is not required.

### Data collection, management and analysis

#### Data collection methods [18a]

The details and schedule of trial assessments (Table 2) are based on the pilot RCT.(28) Assessments take place at baseline (before randomisation), 6 months, 12 months (end of intervention period) and 24 months (12 months after intervention end).

Screening is completed according to the trial eligibility criteria and guided by and documented in a trial screening sheet.

Researchers, trained in data collection processes, collect outcome data during visits to women’s homes (or another venue of the participant’s choosing e.g., University/Community venue). Height, weight and waist circumference are measured by the researcher using standardised protocols and calibrated scales. A hard board is available to place under the scales on uneven surfaces. Baseline demographic information is collected by the researcher. Health service identification number is collected from any woman who provides consent for future data linkage. All remaining outcome data including health economic outcomes are collected via participant self-report questionnaires, either on paper (during the visit or a hard copy left with the participant along with a paid return envelope) or online (link to questionnaire hosted on Qualtrics survey software sent to participant and managed by the trial coordinating team in Queen’s University Belfast (QUB)), based on participant preference at each visit. Researchers offer any required assistance with paper questionnaire completion at the visit or via telephone.

Interviews are conducted with approximately 50 women at each timepoint, with participants purposively sampled to include ethnic, geographical and socio-demographic diversity (i.e., including a spread across trial sites, ethnicity, income level, parity, mental health status, randomised groups and intervention engagement e.g., those who have discontinued messages). Interviews follow a semi-structured interview guide and a translator can be used when any woman would prefer to be interviewed in a language other than English.

Text message engagement data is captured by the LSHTM system, including all messages sent and received organised by participant ID and date.

If the intervention is shown to be effective at 12 months, we will invite a range of relevant stakeholders (e.g., midwives, GPs, PPI, commissioners, policymakers) identified from research team and TSC networks to take part in interviews to explore factors relevant to engagement, scale-up and implementation.

Qualitative data is collected from participants and stakeholders using telephone/Microsoft teams interviews, which are audio-recorded and transcribed.

#### Plans to promote participant retention and complete follow-up [18b]

Home visits are offered for data collection to facilitate women’s participation in research assessments, although visits can take place elsewhere if preferred by the participant. Visits are flexibly arranged for a date and time of convenience for the participant. At the end of each visit, the appointment for the next visit is scheduled if possible. Participants are sent reminders about scheduled visits at one month, one week and one day before the scheduled date and time. We attempt to collect data in a 2-week window either side of the follow-up due date i.e., date ±2 weeks, but allow flexibility outside of this window when women are actively engaging with the research team to arrange the visit. Participants’ preferred methods of contact are documented and prioritised. Three contact attempts using different methods e.g., text, phone, email, are made to all women to confirm scheduled visits and following non-attendance to reschedule. Women who are unable to complete a visit with the researcher at 6 months are sent a link to complete the questionnaire (via Qualtrics email link). As a last resort, when all attempts to arrange a face-to-face visit at 12 (primary endpoint) and 24 months are unsuccessful, participants are asked to provide self-reported weight and complete a questionnaire (via Qualtrics email link). Participants are sent a £25 voucher at each trial timepoint (0, 6, 12 and 24 months; £100 maximum over 24 months) as a token of appreciation for the time committed to completing trial assessments. Participants lost to follow-up at one trial timepoint but who have not withdrawn, are invited to attend future visits.

To maximise data completeness, site researchers follow-up on any non-returned questionnaires and check all received data forms/e-forms to document data queries (e.g., missing data, invalid ranges etc) and any attempts made to resolve them e.g. clarify responses with the participant, using a data query log.

#### Data Management [19]

The hard copy of the CRF which records data at each timepoint including weight is the source data and requires sign off by the completing researcher. For questionnaires, the hard copy or electronic record is considered the source data depending on mode of completion.

The Northern Ireland Clinical Trials Unit (NICTU) are responsible for developing and managing trial databases on validated and secure systems that back-up data, maintain audit trails of data changes and only allow password-protected access to designated trained team members. Screening and CRF data are entered into trial databases by site researchers and a 10% random quality control check of data completed by a different researcher. For CONSORT reporting, screening data includes anonymised summary information for women who do not proceed to randomisation. Paper questionnaires are scanned at site then the original posted to NICTU for data entry directly into the trial database.

Entered data is processed as per the trial specific Data Management Plan. Data queries are generated electronically by NICTU and site staff are required to respond to clarify data, provide missing information or make relevant amendments within the trial databases.

Online questionnaire completion is stored by the Qualtrics system on EU based servers compliant with General Data Protection Regulation (GDPR) and exported by the Trial Manager for merging with the NICTU questionnaire database by the Trial Statistician (CC). Following completion of data collection, data is permanently deleted from Qualtrics.

Qualitative interview transcripts are anonymised and checked for accuracy against the audio recordings, then the recordings deleted.

A minimal amount of personal information (participant’s first name, mobile phone number and weeks postpartum at baseline) is entered into the randomisation system to allow delivery of text messages according to group allocation. Only designated trained team members have password-protected access to the randomisation system.

Text message engagement data is downloaded from the LSHTM system at the end of the intervention period in the form of all messages sent to and replies received from participants.

At the trial end, sites and NICTU send original source documentation (hard copy or electronic) along with consent forms and contact details for individuals who provided consent to be contacted about follow-up to this trial/future-related research/future data linkage to routinely collected health records, to QUB for archiving in accordance with the Sponsor’s requirements (10 years after end of trial/publication). The CI will conduct periodic reviews (every five years) to ensure necessity of data retention. Any other documents containing personal data are confidentially destroyed. Sites are responsible for archiving general site files accumulated during the day-to-day operation of the trial.

### Data protection and confidentiality [27]

All data is collected, stored and handled in accordance with the QUB Research Management Policy (2015), GDPR and any future relevant data protection legislation and all trial team members must be compliant with these regulations. Detailed description of the trial data processing is documented in a Data Privacy Impact Assessment and in the privacy notice given to women expressing an interest in the trial.

Participants are allocated a unique 5-digit participant identifier at randomisation which is used on all data collection forms/files to allow identification of data for each participant, with the key for unlocking pseudonymised data held by site teams in password-protected files. Hard copies of forms are securely stored in locked filing cabinets in locked offices in buildings requiring keypad access and which are alarmed outside normal working hours.

Consent and contact details forms, containing identifiable information, are stored separately to other data (CRFs and questionnaires) and will not be kept for longer than is required. All electronic forms or data files are held on password-protected systems that are routinely backed up and are only accessible using username, password and/or multi-factor authentication. Third-party transcription providers sign a confidentiality agreement detailing data handling requirements. Any file transfer between QUB and other parties (sites/transcription provider) is done, subject to relevant data sharing agreements, via secure and encrypted file transfer systems (e.g., QUB Dropoff). Participants will not be identifiable from any published trial report.

Data collected from women expressing an interest in trial participation but who are not recruited will be confidentially shredded/deleted.

### Access to data [29/31c]

Access to trial records and source data can be granted to authorised Sponsor (or delegate), REC, host institution and regulatory authority representatives for trial-related monitoring, audits and inspections, as per participant consent.

Only employed trial team members have access to personal/sensitive data of participants and data is shared with those responsible for the healthcare of the participants only when health concerns are raised e.g., EPDS score of nine or greater, see [30].

The Trial Manager, Statistician and Health Economist will have access to the anonymised dataset at the trial end, to permit analysis.

The CI will manage access rights to the trial data set, in collaboration with the sponsor, and formal requests for data access need to be made in writing to the CI. The trial will comply with the good practice principles for sharing individual participant data from publicly funded trials and data sharing will be undertaken in accordance with the required regulatory requirements. Any transfer of data to other institutions will be governed by a Data Access Agreement and will be in the form of an anonymised dataset. In the event of publications arising from further data analyses, those responsible will need to provide the CI with a copy of any intended manuscript for approval prior to submission.

### Statistics and analysis

Aligned with best practice and following appropriate reporting guidelines, a Statistical Analysis Plan,(79) Health Economics Analysis Plan (80, 81) and Process Evaluation Analysis Plan are written and approved by the PMT and TSC prior to analysis. Trial results will be reported in accordance with the CONSORT guidelines, with the flow of participants summarised in a CONSORT flow diagram (http://www.consort-statement.org/) and tables of summary baseline characteristics presented, by total recruited sample and 12 month completers.

#### Primary outcome analysis [20a]

Weight change (kg) at 12 months will be compared on an intention to treat basis (i.e., all participants as randomised and regardless of whether or not they engaged with the text messages), between the intervention and active control groups, adjusted for weight at baseline, site, recruitment method (community vs. NHS) and ethnicity. The adjusted difference in means between the groups, corresponding 95% confidence intervals (CIs) and P-value will be reported. A complete-case approach will be used. Sensitivity analysis assessing the impact on the primary outcome analysis will be conducted to: 1) include participant self-reported weight, only where researcher-measured weight is not available; and 2) use last observation carried forward, multiple imputation to impute missing outcome values and delta methods to explore the impact of worse or better outcomes in the individuals with missing data.

The independent TSC in conjunction with NIHR will decide if the trial should progress to stage 3 (24 month follow-up and stakeholder engagement) based on assessment of the 12 month primary outcome data and any other core outcome data required to make a fully informed decision at 12 months (end of intervention). If the trial proceeds, a further analysis of weight change (kg) will be conducted at 24 months. Despite the sequential nature of this comparison, we do not intend to change the significance level because this test will be a secondary analysis.

All testing will be done at the two-sided 5% significance level. All analysis will be conducted using STATA software (Statcorp).

#### Secondary outcome analysis [20a]

Similar analyses will be conducted for secondary continuous outcomes and for secondary analyses of outcomes at other time points. Binary outcomes will be analysed using logistic regression, comparing the intervention and active control groups whilst adjusting for site, recruitment method and ethnicity. Adjusted odds ratios with 95% CIs and corresponding P-values will be reported. Secondary analyses will be considered exploratory and hence the P-values will not be altered to control for multiple testing. The focus of all secondary analyses will be on estimating the difference between-groups with 95% CIs rather than hypothesis testing.

#### Methods for additional analyses (e.g., subgroup analyses) [20b]

To understand any differential effects of the intervention, subgroup analyses will be conducted stratifying by pre-specified variables including site, SES, ethnicity, recruitment method, weeks postpartum (at baseline), BMI (at baseline) and parity. Interaction tests will be conducted, separately, by including interaction terms within regression models. The significance level for subgroup analyses will not be reduced but will be interpreted cautiously. As recommended, all subgroup analyses will be reported.(82)

#### Statistical methods to handle protocol non-adherence and missing data [20c]

The primary analysis of weight change (kg) at 12 months will be on an intention to treat basis.

#### Interim analyses [21b]

No interim analyses are planned.

#### Economic evaluation

The within trial economic evaluation builds on the methods successfully used in the pilot economic evaluation (28) including the identification, measurement and valuation of resource use and expenditure and quality of life/capability wellbeing (Table 2). The economic evaluation is conducted from a UK NHS and personal social services perspective with the addition of a societal perspective applied to capture broader impacts, consistent with the UK’s NICE guidance for public health economic evaluations.(83) Direct costs to the health care system include the intervention implementation costs and any follow-up service use costs. These health and personal perspectives will be assessed to investigate a broader impact of the intervention, including direct cost to participants and indirect costs. Both costs and health outcomes will be discounted at the same annual rate of 1.5% for the reference case, as recommended by the NICE public health methods guidelines.(83) The economic analysis uses a ‘multi-pronged’ approach. The total and mean per-participant costs for the intervention and control groups will be calculated. A sensitivity analysis will incorporate development costs. Regression analysis, controlling for baseline differences, will be used to estimate the average cost per-participant and the average quality-adjusted life year (QALY) achieved, by group. A cost-utility analysis will report cost per QALY gained over the 24 month period using the area under the curve approach.(84) Results will be presented on the cost-effectiveness plan and 95% CIs for the incremental cost-effectiveness ratio will be determined.(85) Joint uncertainty in costs and outcomes will be represented using a cost-effectiveness acceptability curve to present the probability of the SMS intervention being cost-effective for prevailing UK ceiling thresholds for costs per QALY gained.(86) The cost-effectiveness analysis will align with the primary outcome, by examining differences in weight and BMI and total costs between treatment groups, the incremental cost per weight gain averted will be calculated. The mean costs and effects for each trial arm will be calculated and presented along with the incremental costs and effects between arms (including 95% CIs).

The within trial economic analysis will be carried out for the 24 months follow-up and will form the primary analysis. However, this short time horizon may be insufficient to capture the total costs and benefits related to the intervention. If differences in quality of life or capability wellbeing are identified, the results will be extrapolated to a lifetime horizon using modelling methods.

#### Monitoring and auditing [23]

As per contractual agreements and participant consent, direct access to trial records and source documents will be granted to the Sponsor or regulatory agencies by all research staff and trial sites, for the purposes of trial related monitoring, audits and inspections. The trial is monitored and audited in accordance with the Sponsor’s policy, which is consistent with the UK Policy Framework for Health and Social Care Research, and the TSC informed of the main findings of any monitoring, audit or inspection.

#### Trial Steering Committee (TSC) and Data Monitoring [5d/21a]

The independent TSC meet face-to-face or online at least annually, as required, to provide trial oversight on behalf of the sponsor and funder. The TSC determine trial progression at the recruitment mid-point (see Figure 1 and [15]) and primary outcome stage. As this RCT involves a low-risk behaviour change lifestyle intervention which is unlikely to raise concerns for participant safety (no evidence of harm was reported in the pilot study or in any other text message behaviour change studies), the sponsor and funder agreed that a Data Monitoring Committee would not be convened [21a]. Participant safety is a standing agenda item at each meeting, see [22].

### Project Management Team (PMT) [5d]

The PMT is the key decision-making group consisting of the co-applicants, Trial Manager and PPI representative. The PMT meet online every 2-4 months to oversee the trial design, management and conduct, including monitoring participant safety, see [22].

#### Trial coordinating centre [5d]

The trial is coordinated by the Chief Investigator (MMcK) and the Trial Manager (DG) in QUB. Weekly meetings between the CI and Trial Manager and regular meetings with site teams are held, to discuss trial activities across the five trial sites. Site PIs are responsible for managing staff and overseeing participant recruitment and follow-up at their sites.

### Adverse event reporting and harms [22]

Serious adverse events (SAEs) are recorded by researchers from the time of the participant’s consent into the trial until one month after the end of the 12 month intervention period and reported in line with GCP guidelines and the Sponsor’s adverse event reporting procedures. Site PIs review SAEs within 24 hours to determine severity, relatedness and expectedness. The CI (or delegate) is informed of any event evaluated as being severe, related and unexpected, to conduct a second review. Onward reporting of any SAEs categorised as related and expected by both the PI and CI is actioned by the CI or Trial Manager to the Sponsor and REC within 24 hours and 15 days respectively. All SAEs are summarised and included in annual reports to the REC and reported during meetings of the PMT and TSC.

### Ancillary and post-trial care [30]

This is deemed a low-risk public health trial and no evidence of harm has been reported in text message studies for other health behaviours or in our pilot RCT, so no post-trial care is delivered. All women are given a signposting ‘Useful Contacts’ leaflet during trial visits advising them on relevant agencies and organisations that they might contact for support on a variety of issues e.g., bereavement. We monitor any emerging safety issues or unintended consequences and any participants experiencing such issues will be referred to their GP for appropriate treatment. This includes the onward reporting of EPDS scores of nine or above (Table 2). The EPDS is a screening questionnaire used to detect postnatal depressive symptoms and the scale guidance indicates that women with scores meeting this threshold would benefit from further clinical assessment. We calculate scores within one week of receiving the completed EPDS at each follow-up timepoint and letters are sent to the GP and the participant to inform of scores ≥9, as per PIS and consent processes.

#### Personal and Public Involvement (PPI)* Statement

*(* Note: In NI, PPI means Personal and Public Involvement due to integrated Health and Social Care systems. The rest of the UK use ‘Patient and Public Involvement’ also known as ‘Patient and Public Involvement and Engagement’)*

PPI was integral to the pilot study with guidance received on the development of the text message libraries and all trial methods and materials, as previously reported.(28) The design of the present trial mirrors the methods used in the pilot trial. We recruited PPI representatives from Scotland, London and Bradford to conduct a period of trial adaptation (Figure 1) whereby the library of intervention text messages and trial recruitment materials underwent further PPI to ensure content was acceptable and culturally relevant for women from a range of backgrounds and ethnicities across the UK.

We have PPI representatives on our PMT and independent TSC to contribute to decision making throughout the trial and we seek their advice on an ad-hoc basis, as matters arise. PPI representatives will help shape the dissemination plan for trial results and will review dissemination materials to ensure communication of findings in an appropriate manner.

PPI work is led by the trial team and conducted and reimbursed according to NIHR guidance for involving the public in research.(87)

#### Protocol amendments [25]

The CI, in consultation with the PMT and Sponsor, is responsible for the decision to amend the protocol and, in turn, for communicating substantive changes to relevant stakeholders. Substantial and non-substantial amendments are reviewed by the sponsor to determine the appropriate amendment category then submitted to the approving REC via the IRAS system for approval prior to implementation, and subsequently disseminated to participating centres (R&D offices, regulatory agencies). Version control and details of amendment history are clearly documented in the protocol and are disseminated to the trial sites, PMT and TSC members, funder and trial registries.

The current protocol is 3.0 (10/05/2023). Amendments to the protocol incorporated: 1) the added mental health objectives resulting from the additional NIHR funding; 2) an increase in trial sample size (from 850 to 888 women) to allow for higher estimated pregnancy exclusion rates in more ethnically diverse populations than were originally accounted for; 3) to include processes to collect participant self-reported weight at 12 and 24 months only as a last resort in cases where it is not possible to obtain researcher-measured weight; and 4) to offer a booklet summarising the intervention content to the active control group after completion of data collection at 24 months.

#### Protocol training and deviations

All trial staff undergo training on the protocol, trial processes and standard operating procedures (SOPs) for which they are responsible (detailed on the site delegation log). The Trial Manager monitors trial activities across sites to ensure adherence and consistency to the protocol. There are no planned protocol deviations. If a deviation occurs, it is documented and reported immediately to the CI and Sponsor, including corrective and preventative actions. Frequent recurrence of protocol deviations is unacceptable, requires immediate action and could be classified as a serious breach i.e., one which is likely to significantly affect the safety or physical or mental integrity of trial participants and/or the scientific value of the trial. Any breach meeting this definition is immediately reported to the Sponsor, as per the Sponsor’s SOP on ‘Matters of non-compliance with study protocol’.

#### Peer review

This trial has undergone independent, expert peer-review as part of the NIHR PHR funding process. It has also been reviewed and approved by the QUB Research Governance, Ethics and Integrity Manager, on behalf of the Sponsor.

## Ethics and dissemination

### Ethical and Regulatory considerations [24]

The trial is conducted in accordance with the recommendations for physicians involved in research on human participants adopted by the 18^th^ World Medical Assembly, Helsinki 1964, and later revisions, the UK Policy Framework for Health and Social Care, relevant UK data protection legislation and the principles of GCP. QUB acts as sponsor for the trial and has in place comprehensive SOPs for the approval and monitoring of research. This protocol and other relevant trial documents were reviewed and approved by the West of Scotland Research Ethics Service REC 4 22/WS/0003. Correspondence with the REC is retained in the Trial Master File. An annual progress report is submitted to REC until the trial end. The CI will report to REC when the trial ends (or is terminated prematurely) and a final report with trial results submitted within one year of this.

Appropriate approvals from participating organisations e.g., NHS R&D approvals are obtained before recruiting via applicable routes.

### Dissemination [31a]

Dissemination plans informed by PPI and participant and stakeholder engagement will be developed and reviewed annually. A range of outputs including NIHR final report, peer-reviewed open access publications, conference presentations, policy briefings, non-academic research summaries and a draft scaling-up plan, are anticipated to be disseminated relevant for different audiences, including trial participants, postpartum women throughout the UK, healthcare professionals, service commissioners, academics, the Government and public health bodies.

## Supporting information

SPIRIT checklist

TIDieR checklist

## Data Availability

All data produced in the present study are available upon reasonable request to the authors

## Authors’ contributions [31b]

MCM led the funding proposal and protocol development with support from all co-authors (DG, ES, ASA, SB, CC, EC, SUD, CF, SH, PH, FK, CMcD, EM, JVW) who provided expert input on trial design and intervention content. MCM, ES and DG co-ordinated the ethical approval. DG drafted the protocol manuscript. All authors (DG, ES, ASA, SB, CC, EC, SUD, CF, SH, PH, FK, CMcD, EM, JVW, MCM) contributed to the revision of the manuscript and have read and approved the final version.

## Funding Statement [4]

This trial is supported by the NIHR Public Health Research (PHR) Programme (NIHR131509) and intervention costs provided by the Northern Ireland Public Health Agency.

## Competing interests’ statement [28]

The CI, PIs, co-investigators, trial staff and TSC are required to disclose information on competing interests that might influence the trial integrity, such as commercial ownership interests. A record of competing interests is retained by the Trial Manager for reporting in all trial publications.

## Acknowledgements

For the purpose of open access, the author has applied a Creative Commons Attribution (CC BY) licence to any Author Accepted Manuscript version arising from this submission.

## Notes

### Competing Interest Statement

The authors have declared no competing interest.

### Clinical Trial

ISRCTN16299220

### Author Declarations

REC 4 of West of Scotland Research Ethics Service gave ethical approval for this work (reference 22/WS/0003).

