## Supplemental Appendices for "Effectiveness and cost effectiveness of a 12 month automated text message intervention for weight management in postpartum women with overweight or obesity: protocol for the Supporting MumS (SMS) multi-site, parallel-group, randomised controlled trial"

**Supplemental appendix 1- SPIRIT checklist (2)**

SPIRIT 2013 Checklist: Recommended items to address in a clinical trial protocol and related documents*

| **Section/item** | **Item No** | **Description** | **Addressed on page number** |
| --- | --- | --- | --- |
| **Administrative information** | | |  |
| Title | 1 | Descriptive title identifying the study design, population, interventions, and, if applicable, trial acronym | 4 |
| Trial registration | 2a | Trial identifier and registry name. If not yet registered, name of intended registry | 2 |
|  | 2b | All items from the World Health Organization Trial Registration Data Set | Supplemental appendix 5 |
| Protocol version | 3 | Date and version identifier | 4 |
| Funding | 4 | Sources and types of financial, material, and other support | 50 |
| Roles and responsibilities | 5a | Names, affiliations, and roles of protocol contributors | 4-7, 49-50 |
|  | 5b | Name and contact information for the trial sponsor | 7 |
|  | 5c | Role of study sponsor and funders, if any, in study design; collection, management, analysis, and interpretation of data; writing of the report; and the decision to submit the report for publication, including whether they will have ultimate authority over any of these activities | 7 |
|  | 5d | Composition, roles, and responsibilities of the coordinating centre, steering committee, endpoint adjudication committee, data management team, and other individuals or groups overseeing the trial, if applicable (see Item 21a for data monitoring committee) | 40 |
| **Introduction** |  |  |  |
| Background and rationale | 6a | Description of research question and justification for undertaking the trial, including summary of relevant studies (published and unpublished) examining benefits and harms for each intervention | 8-12 |
|  | 6b | Explanation for choice of comparators | 12-13 |
| Objectives | 7 | Specific objectives or hypotheses | 13-14 |
| Trial design | 8 | Description of trial design including type of trial (eg, parallel group, crossover, factorial, single group), allocation ratio, and framework (eg, superiority, equivalence, noninferiority, exploratory) | 14 |
| **Methods: Participants, interventions, and outcomes** | | |  |
| Study setting | 9 | Description of study settings (eg, community clinic, academic hospital) and list of countries where data will be collected. Reference to where list of study sites can be obtained | 14 |
| Eligibility criteria | 10 | Inclusion and exclusion criteria for participants. If applicable, eligibility criteria for study centres and individuals who will perform the interventions (eg, surgeons, psychotherapists) | 14-15, 26-27 |
| Interventions | 11a | Interventions for each group with sufficient detail to allow replication, including how and when they will be administered | 15-19 |
|  | 11b | Criteria for discontinuing or modifying allocated interventions for a given trial participant (eg, drug dose change in response to harms, participant request, or improving/worsening disease) | 19 |
|  | 11c | Strategies to improve adherence to intervention protocols, and any procedures for monitoring adherence (eg, drug tablet return, laboratory tests) | 19 |
|  | 11d | Relevant concomitant care and interventions that are permitted or prohibited during the trial | 19-20 |
| Outcomes | 12 | Primary, secondary, and other outcomes, including the specific measurement variable (eg, systolic blood pressure), analysis metric (eg, change from baseline, final value, time to event), method of aggregation (eg, median, proportion), and time point for each outcome. Explanation of the clinical relevance of chosen efficacy and harm outcomes is strongly recommended | 20-23 |
| Participant timeline | 13 | Time schedule of enrolment, interventions (including any run-ins and washouts), assessments, and visits for participants. A schematic diagram is highly recommended (see Figure) | 23-24 |
| Sample size | 14 | Estimated number of participants needed to achieve study objectives and how it was determined, including clinical and statistical assumptions supporting any sample size calculations | 25-26 |
| Recruitment | 15 | Strategies for achieving adequate participant enrolment to reach target sample size | 26 |
| **Methods: Assignment of interventions (for controlled trials)** | | |  |
| Allocation: |  |  |  |
| Sequence generation | 16a | Method of generating the allocation sequence (eg, computer-generated random numbers), and list of any factors for stratification. To reduce predictability of a random sequence, details of any planned restriction (eg, blocking) should be provided in a separate document that is unavailable to those who enrol participants or assign interventions | 28-29 |
| Allocation concealment mechanism | 16b | Mechanism of implementing the allocation sequence (eg, central telephone; sequentially numbered, opaque, sealed envelopes), describing any steps to conceal the sequence until interventions are assigned | 29 |
| Implementation | 16c | Who will generate the allocation sequence, who will enrol participants, and who will assign participants to interventions | 29 |
| Blinding (masking) | 17a | Who will be blinded after assignment to interventions (eg, trial participants, care providers, outcome assessors, data analysts), and how | 29-30 |
|  | 17b | If blinded, circumstances under which unblinding is permissible, and procedure for revealing a participant’s allocated intervention during the trial | 30 |
| **Methods: Data collection, management, and analysis** | | |  |
| Data collection methods | 18a | Plans for assessment and collection of outcome, baseline, and other trial data, including any related processes to promote data quality (eg, duplicate measurements, training of assessors) and a description of study instruments (eg, questionnaires, laboratory tests) along with their reliability and validity, if known. Reference to where data collection forms can be found, if not in the protocol | 30-31 |
|  | 18b | Plans to promote participant retention and complete follow-up, including list of any outcome data to be collected for participants who discontinue or deviate from intervention protocols | 31-32 |
| Data management | 19 | Plans for data entry, coding, security, and storage, including any related processes to promote data quality (eg, double data entry; range checks for data values). Reference to where details of data management procedures can be found, if not in the protocol | 32-34 |
| Statistical methods | 20a | Statistical methods for analysing primary and secondary outcomes. Reference to where other details of the statistical analysis plan can be found, if not in the protocol | 36-37 |
|  | 20b | Methods for any additional analyses (eg, subgroup and adjusted analyses) | 37-38 |
|  | 20c | Definition of analysis population relating to protocol non-adherence (eg, as randomised analysis), and any statistical methods to handle missing data (eg, multiple imputation) | 38 |
| **Methods: Monitoring** | | |  |
| Data monitoring | 21a | Composition of data monitoring committee (DMC); summary of its role and reporting structure; statement of whether it is independent from the sponsor and competing interests; and reference to where further details about its charter can be found, if not in the protocol. Alternatively, an explanation of why a DMC is not needed | 40 |
|  | 21b | Description of any interim analyses and stopping guidelines, including who will have access to these interim results and make the final decision to terminate the trial | 38 |
| Harms | 22 | Plans for collecting, assessing, reporting, and managing solicited and spontaneously reported adverse events and other unintended effects of trial interventions or trial conduct | 41 |
| Auditing | 23 | Frequency and procedures for auditing trial conduct, if any, and whether the process will be independent from investigators and the sponsor | 39-40 |
| **Ethics and dissemination** | | |  |
| Research ethics approval | 24 | Plans for seeking research ethics committee/institutional review board (REC/IRB) approval | 2, 44 |
| Protocol amendments | 25 | Plans for communicating important protocol modifications (eg, changes to eligibility criteria, outcomes, analyses) to relevant parties (eg, investigators, REC/IRBs, trial participants, trial registries, journals, regulators) | 42-44 |
| Consent or assent | 26a | Who will obtain informed consent or assent from potential trial participants or authorised surrogates, and how (see Item 32) | 27-28 |
|  | 26b | Additional consent provisions for collection and use of participant data and biological specimens in ancillary studies, if applicable | 28 |
| Confidentiality | 27 | How personal information about potential and enrolled participants will be collected, shared, and maintained in order to protect confidentiality before, during, and after the trial | 34-35 |
| Declaration of interests | 28 | Financial and other competing interests for principal investigators for the overall trial and each study site | 50 |
| Access to data | 29 | Statement of who will have access to the final trial dataset, and disclosure of contractual agreements that limit such access for investigators | 35 |
| Ancillary and post-trial care | 30 | Provisions, if any, for ancillary and post-trial care, and for compensation to those who suffer harm from trial participation | 41-42 |
| Dissemination policy | 31a | Plans for investigators and sponsor to communicate trial results to participants, healthcare professionals, the public, and other relevant groups (eg, via publication, reporting in results databases, or other data sharing arrangements), including any publication restrictions | 44-45 |
|  | 31b | Authorship eligibility guidelines and any intended use of professional writers | 49-50 |
|  | 31c | Plans, if any, for granting public access to the full protocol, participant-level dataset, and statistical code | 35 |
| **Appendices** |  |  |  |
| Informed consent materials | 32 | Model consent form and other related documentation given to participants and authorised surrogates | Supplemental appendix 4 |
| Biological specimens | 33 | Plans for collection, laboratory evaluation, and storage of biological specimens for genetic or molecular analysis in the current trial and for future use in ancillary studies, if applicable | N/A |

**Supplemental appendix 2- TIDieR checklist (52)**

| **Item number** | **Item** | **Where located **** | |
| --- | --- | --- | --- |
|  |  | Primary paper  (page or appendix  number) | Other ^†^ (details) |
|  | **BRIEF NAME** |  |  |
| **1.** | Provide the name or a phrase that describes the intervention. | 15 |  |
|  | **WHY** |  |  |
| **2.** | Describe any rationale, theory, or goal of the elements essential to the intervention. | 15-16 and Supplemental appendix 3 | <https://doi.org/10.3310/phr08040> (28) |
|  | **WHAT** |  |  |
| **3.** | Materials: Describe any physical or informational materials used in the intervention, including those provided to participants or used in intervention delivery or in training of intervention providers. Provide information on where the materials can be accessed (e.g. online appendix, URL). | 16-17 and Supplemental appendix 3 | _____________ |
| **4.** | Procedures: Describe each of the procedures, activities, and/or processes used in the intervention, including any enabling or support activities. | 16-17 | _____________ |
|  | **WHO PROVIDED** |  |  |
| **5.** | For each category of intervention provider (e.g. psychologist, nursing assistant), describe their expertise, background and any specific training given. | 17 | _____________ |
|  | **HOW** |  |  |
| **6.** | Describe the modes of delivery (e.g. face-to-face or by some other mechanism, such as internet or telephone) of the intervention and whether it was provided individually or in a group. | 17 | _____________ |
|  | **WHERE** |  |  |
| **7.** | Describe the type(s) of location(s) where the intervention occurred, including any necessary infrastructure or relevant features. | 17 | _____________ |
|  | **WHEN and HOW MUCH** |  |  |
| **8.** | Describe the number of times the intervention was delivered and over what period of time including the number of sessions, their schedule, and their duration, intensity or dose. | 18 | <https://doi.org/10.3310/phr08040> (28) |
|  | **TAILORING** |  |  |
| **9.** | If the intervention was planned to be personalised, titrated or adapted, then describe what, why, when, and how. | 15-18 | _____________ |
|  | **MODIFICATIONS** |  |  |
| **10.^ǂ^** | If the intervention was modified during the course of the study, describe the changes (what, why, when, and how). | N/A | _____________ |
|  | **HOW WELL** |  |  |
| **11.** | Planned: If intervention adherence or fidelity was assessed, describe how and by whom, and if any strategies were used to maintain or improve fidelity, describe them. | 19 | _____________ |
| **12.^ǂ^** | Actual: If intervention adherence or fidelity was assessed, describe the extent to which the intervention was delivered as planned. | N/A | _____________ |

** **Authors** - use N/A if an item is not applicable for the intervention being described. **Reviewers** – use ‘?’ if information about the element is not reported/not sufficiently reported.

† If the information is not provided in the primary paper, give details of where this information is available. This may include locations such as a published protocol or other published papers (provide citation details) or a website (provide the URL).

ǂ If completing the TIDieR checklist for a protocol, these items are not relevant to the protocol and cannot be described until the study is complete.

**Supplemental appendix 3- SMS Logic Model and sample text messages**

a) **Supporting MumS intervention logic model**

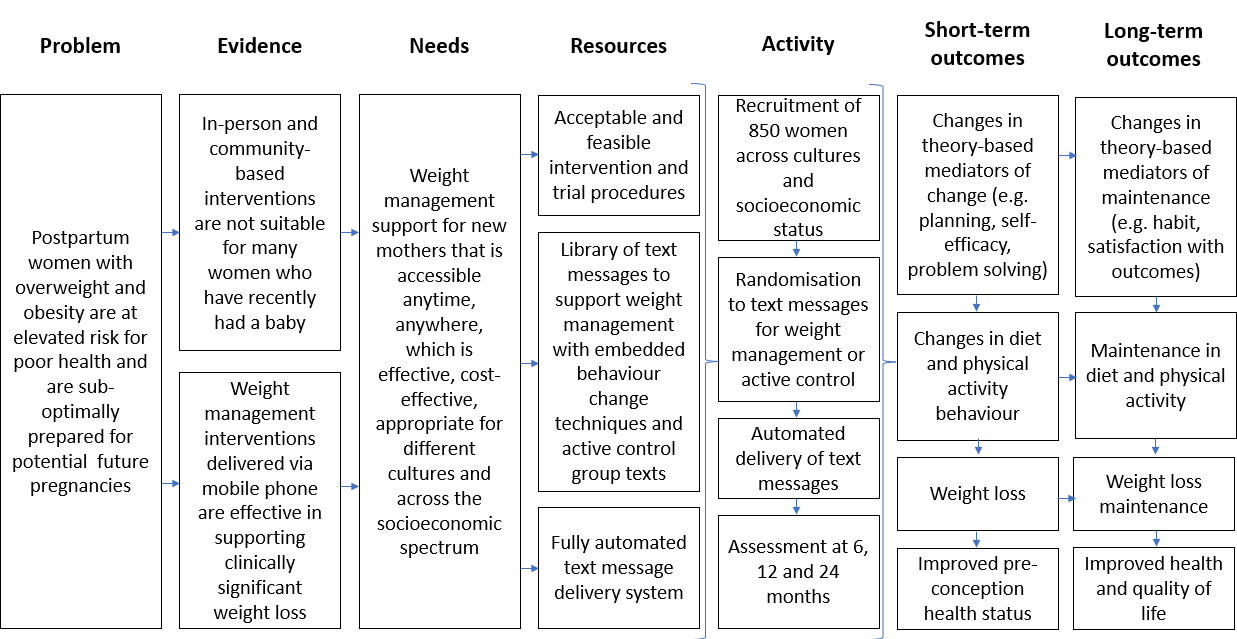

b) **Supporting MumS sample text messages**

| **Type of text message** | | **Example of message sent** | **Example of automated responses (if applicable)** |
| --- | --- | --- | --- |
| **Core intervention text message** | | *‘Another tip for success: Keep track of what you eat. People who regularly monitor what they eat are more likely to meet their weight loss target. Stay tuned for more texts on food tracking’.* |  |
| **Weekly self-weighing reminder** | **Months 0-6** | *‘What are the scales saying this week? Text us e.g. 11 st 5, or 73.3 kg’.* | N/A |
|  | **Months 7-12** | *‘Weekly weighing = your very own early warning system! Step on the scales today and text us your weight followed by UP, DOWN, SAME. e.g. “10 st 7 SAME” or “66.7 kg SAME’.* | Up: *‘Don't give up! The best thing to do is prevent these small regains from turning into bigger regains. What could you do this week? E.g. maybe exercise a little more or be more mindful of portion sizes?’* |
|  |  |  | Down: *‘Success! Your efforts are paying off! All the hard work is worth it :)’.* |
|  |  |  | Same: *‘Maintaining your weight is something to be proud of! Treat yourself to a reward e.g. a relaxing bath or simply 10 minutes to yourself!’* |
| **Yes/No question** | | ‘*WEEK 14. Have you set your activity goal for the week ahead? Text Y11 for yes or N11 for no’.* | Y11 (yes): ‘*Great job, setting goals is clearly becoming one of your weekly habits :) Alongside your activity goal, set a food goal too and you're onto a double winner!’* |
|  |  |  | N11 (no): ‘*New to the whole activity thing? Set a goal to do a small amount of an activity that you enjoy - for example, a brisk 15-minute walk or a zumba routine when your little one goes to sleep. It will help motivate you and give you a boost of confidence to do more next time’*. |
| **Trigger words** | | *‘You can always text us back for more help – text us with one of these trigger words and texts with instant support will appear on your phone: ‘BAD DAY’, ‘SLIP-UP’, ‘TIRED’ and ‘CRAVE’.* | Bad day*: ‘Everyone has these type of days. Don't be hard on yourself! Press delete for the day so far and start afresh!’* |
|  |  |  | Slip-up*: ‘Don't let one slip-up discourage you. You have the ability to pick up right where you left off!’* |
|  |  |  | Tired response: ‘*Fresh air can work wonders! Getting out of the house for a while can help you think more clearly and boost your energy’.* |
|  |  |  | Crave response: *‘Tried brushing your teeth or chewing a piece of sugar free gum? It might work!’* |
| **Weight management when breastfeeding** | | *‘Breastfeeding can make you hungry. Just fill up on the good stuff (fruit and vegetables, wholegrain cereals) and the good eating habits will stay with you even when the breastfeeding stops :) ‘* | N/A |
| **Smoking cessation and weight management** | | *‘It is only normal to have cravings! Why not exercise to combat it? Try a short walk (even for just 5 mins) if a craving hits, the distraction can really help.’* | N/A |
| **Intervention “buddy system”** | | *‘Welcome to WEEK 34. Many people that have lost weight and kept it off have done so with support from family and friends! You can opt in to get your friend/partner/family member to receive SMS texts for extra support - text 'SUPPORT' followed by the person's mobile number to sign up (ask their permission first). E.g. Support 0780000000’.* | N/A |
| **Active control** | | ***‘****1, 2, 3! Your toddler might know how to count to 3 or more...or they might get some of the numbers mixed up. Whatever stage they are at, just keep encouraging them by counting from 1 to 10’.* | N/A |

**Supplemental appendix 4- Informed consent materials [32]**

a) **Participant Information Sheet**

| 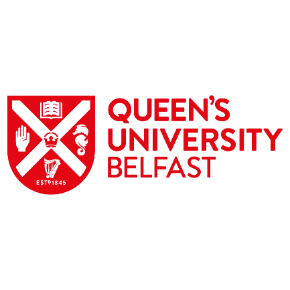 | 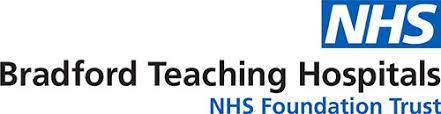 | 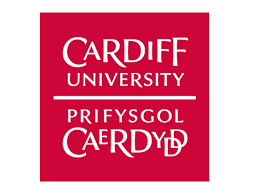 | 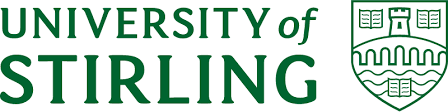 | 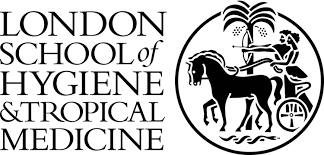 |
| --- | --- | --- | --- | --- |

**PARTICIPANT INFORMATION SHEET**

**The Supporting MumS (SMS) study**

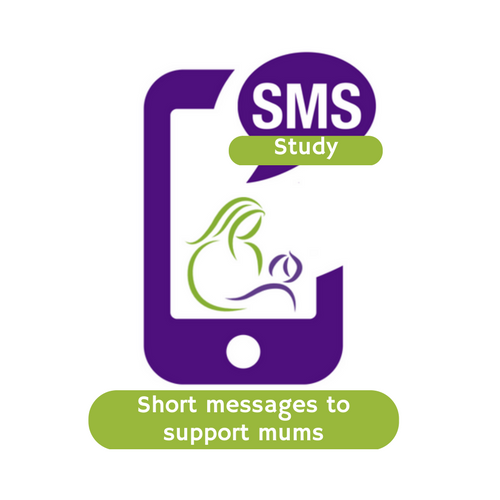

We would like to invite you to take part in this research study which is taking place in Northern Ireland, England, Scotland and Wales.

Before you decide whether to take part, it is important that you understand why we are doing this research and what it will involve.

Please take time to read the information carefully and discuss it with friends/relatives, if you wish. If you have any questions, please contact us using the details given at the end of the sheet.

**Why are we doing this study?**

Welcoming a baby brings lots of joy and happiness but being a new mum also means life is a bit busy! There may be areas where mums could benefit from more support. Mums often talk about having a baby as a time when their weight started to creep up and how they would welcome support with this. We are interested in finding out if text messages can help support weight management in the first two years after having a baby.

*For this study, the term ‘mum’ or ‘mother’ includes people who do not identify as women but are pregnant or have given birth. The term ‘your child’ means your youngest child at the time of signing up to the study.*

**Why have I been asked to take part?**

You have been chosen to take part, as you are over 18 years old and have had a baby within the past two years and have a body mass index over 25kg/m^2^.

**Do I have to take part?**

No, it is up to you whether or not you want to take part. If you do decide to take part, you are free to leave the study at any stage without giving a reason for doing so. Your usual health care will not be affected at any time.

**What will happen if I take part?**

After you read this leaflet we will contact you to discuss the study in more detail and answer any further questions you may have.

If you decide to take part, you will be asked to sign the consent form for the study and complete some study measurements (described below). You will then receive text messages to your mobile phone for 12 months. The number of text messages you receive each week will vary between 3 and 14 per week. The text messages are delivered to you by a secure text message system owned and operated by the London School of Hygiene and Tropical Medicine.

Mums who take part in the study will get text messages on **ONE** of the two topics shown below. It is important that you understand that the topic is randomly chosen for you and that there is a 50:50 chance you will get texts on either topic 1 or topic 2:

1. Information and advice on making healthy food choices and keeping physically active to help you lose weight.

**OR**

2. Information and advice on your child’s health and development.

**What information will you collect from me if I take part?**

We would meet with you 4 times (during a 24 month period) to collect some measurements and information. These visits will take place at the start of the study, at 6 months, at 12 months (when the text messages stop) and at 24 months (12 months after you stopped receiving the text messages).

Start of study

End of study

Visit 1: 0 months

Visit 2: 6 months

Visit 3: 12 months

Visit 4: 24 months

For the visits, the researcher will either visit you at your home or you can come and meet the researcher at a convenient location such as a University building or a local community venue; whichever is most convenient for you.

You will receive a £25 voucher on completion of each visit in recognition of your time to complete the research measures. This will be a total of £100 over the 24-month study, if all visits are completed. Each visit will last approximately 1 hour.

At each visit we will collect the following information:

a) We will measure your height (only at visit 1) and will ask you to step on the scales to collect information on weight and will measure your waist circumference (this will take about 10 minutes).
b) We will ask you to complete a questionnaire booklet about your physical and mental health, lifestyle and wellbeing. The booklet will take approximately 35 minutes to complete. You will have the option of:
   - completing a printed questionnaire booklet in your own time and posting it back to us in a stamped addressed envelope we provide; OR
   - completing the questionnaire online by accessing a link we give you; OR
   - completing the questionnaire with the researcher at the study visit or over the phone.

When (a) and (b) above has been collected from you, we will arrange for the £25 voucher to be sent to you.

We would also ask you to consider taking part in a short (approximately 20 minutes) telephone interview twice during the study (at the 6 month visit and the 12 month visit) to help us understand how you found the text messages. *This part is optional.*

**Linking to routine health-related data in the future (this part is optional)**

We would also ask you to consider allowing us to collect health-related data about you and your child in the future. We would do this by linking with organisations that collect data about the services they provide to women and children. For example, the NHS keeps records, known as ‘routine data’ about how we use different services such as the health service.

Routine data is stored electronically on different systems. To obtain your routine data from other systems, we would share some personally-identifiable details such as name, date of birth, address and NHS number to the organisations that manage these systems and they will send us data back. This will all be done using secure data transfer systems that have been set up carefully to keep your information safe.

Doing this helps us to understand if a study like this has any longer-term benefits and how we might improve health services in the future.

We may access routine data from these sources:

- Health records such as GP and dental records, maternity and health visiting records and disease registers
- Local authority and social care
- Family/children’s centres
- Education or school
- National child measurement programme
- Voluntary organisations

For example, from maternity records we may collect information on any further pregnancies you have and from GP records we may collect information on any diagnosed conditions such as high blood pressure or type 2 diabetes.

If you consent to us linking to routine health-related data on the consent form, we will ask you for your NHS number. This number will only be stored by QUB for 15 years and will only be used for this purpose. It will be stored securely, separately from your other data, in a password protected, encrypted file on computers that are only accessible to authorised members of the research team. If at any time you want us to remove your NHS number from our records, you can do so by contacting us as per details given at the end of this information sheet. Otherwise, it will be removed from our records after 15 years. An example of how we would link your routine data is shown below.

*You do not have to consent to data linkage to participate in this study. This part is optional.*

We provide the data holder, e.g. the NHS Maternity record system, with your NHS number which is unique to you

The NHS matches your NHS number with their records and provides us with your routine maternity data

We add the maternity data to the data we have already collected from you in the SMS study

Example of routine data linkage process

**What are the possible advantages and disadvantages of taking part?**

By taking part in this research you will be helping us find the best ways to support new mums. In the study questionnaires and interviews, we will collect information about your postnatal physical and mental health. This information will be used to help us understand how we might improve services for women after they have a baby. Some of the messages you receive may be helpful for you and/or your baby by helping you to lose weight or by providing you with information about child health and development. We do not anticipate any risks from taking part in this study.

**How will we use information about you?**

We will need to use information from you for this research project. This information will include your name, address, NHS number, date of birth, telephone number, email address and date of birth of your youngest child. People will use this information to do the research or to check your records to make sure that the research is being done properly.

People who do not need to know who you are will not be able to see your name or contact details. Your data will have a code number instead.

We will keep all information about you safe and secure.

Once we have finished the study, we will keep some of the data so we can check the results. We will write our reports in a way that no-one can work out that you took part in the study.

**Will my taking part in the study be kept confidential?**

**Yes,** all your data will be treated with the strictest confidence and your details will not be shared with anybody outside of the research team, unless there is a serious risk of harm to you or others.

Any information collected from you will be stored securely on password protected files on password protected computers that only the research team can access. Hard copies of documents will be kept in locked filing cabinets in locked offices that are only accessible to the research team and are located in a building that is locked outside normal working hours.

Any interviews conducted over the telephone will be audio-recorded and the recording will then be typed up for research purposes. The typed transcript will not contain any names and will be labelled with your unique study number. The recording will be destroyed once the typed transcript is prepared.

Study data will be kept separate from personal information (such as name and address). Only members of the research team will have access to view identifiable data. However, in some instances, inspectors from regulatory authorities may need to access data for checking the quality of the research. All members of the research team and regulatory bodies are trained in data protection and will comply with the requirements of data protection legislation.

Once the study is complete and it is no longer necessary to keep identifiable information or contact details, we will destroy our records of this personal information.

The study questionnaire asks some questions about mental and physical health. If we have any concerns that your responses to such questions may indicate you are at risk of postpartum depression, we will inform you and your GP by letter as part of our duty of care to you. If at any stage we have concerns that you, or someone else, is at risk of harm then we are obliged to tell the relevant services, for example your GP or social services.

**What are your choices about how your information is used?**

- You can stop being part of the study at any time, without giving a reason, but we will keep information about you that we already have.
- We need to manage your records in specific ways for the research to be reliable. This means that we won’t be able to let you see or change the data we hold about you.
- If you agree to take part in this study, you will have the option to take part in future research using your data saved from this study.

**Where can you find out more about how your information is used?**

You can find out more about how we use your information:

- at [www.hra.nhs.uk/information-about-patients/](https://www.hra.nhs.uk/information-about-patients/)
- in our privacy notice from [go.qub.ac.uk/SMSstudy](http://www.go.qub.ac.uk/SMSstudy) or ask the research team for a copy
- by asking one of the research team
- by ringing us on 07341 888415.

**How will I find out the results of this project?**

When the study is finished and the study information has been analysed, we will send you a summary of the results.

**Who is organising and funding the research?**

For this study we will be recruiting women from all four countries in the UK. The work is being led by Professor Michelle McKinley from Queen’s University Belfast. Researchers from Universities in London, Cardiff, Stirling and the Bradford Teaching Hospitals NHS Foundation Trust are leading recruitment in their areas.

Mums have helped us design our text messages and advise on how the study is conducted and will continue to do so.

The study is funded by National Institute of Health Research and the Public Health Agency Northern Ireland.

**Has this study been approved for safety by an ethics committee? Is this study safe?**

This study has been reviewed and approved by the West of Scotland Research Ethics Committee 4 and IRAS ID 305557.

**What will happen if I don’t want to carry on with the study?**

You can withdraw from the study at any time, without giving a reason by contacting the research team (contact details at the end of this leaflet). If you do decide to withdraw from the study, we will use the data collected up to that point, but we will not collect any more data.

**What if something goes wrong?**

There are no special compensation arrangements. Queen’s University Belfast will provide indemnity for this study. If you are harmed due to someone’s negligence, then you may have grounds for legal action but you may have to pay for it.

**What if there is a problem?**

If you have a concern about any aspect of this study, you can speak to the researchers who will do their best to answer your questions (contact details on the last page).

Should you remain unhappy and wish to make a formal complaint, you can contact:

Research Governance Team at Queen’s University Belfast,

63 University Road, Belfast

BT7 1NF

**Contact for Further Information:**

| Researchers’ Name, Address, Email address, Telephone (site specific) | Dr Dunla Gallagher  SMS Trial Manager  Centre for Public Health  Queen's University Belfast School of Medicine, Dentistry & Biomedical Sciences  Institute of Clinical Sciences (Block B)  Grosvenor Road Belfast, N.Ireland BT12 6BA  Tel: 07341 888415  |
| --- | --- |

**Please ask us if there is anything that is not clear or if you would like more information.**

b) **Privacy Notice**

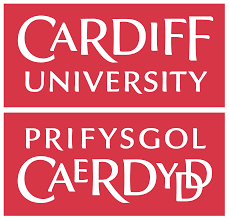

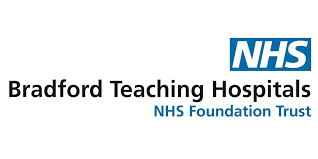

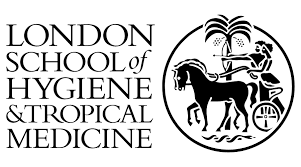

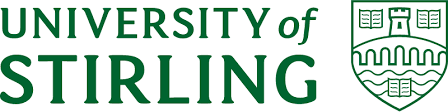

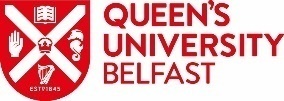

**PRIVACY NOTICE**

**The Supporting MumS (SMS) study**

Queen’s University Belfast, Bradford Teaching Hospitals NHS Foundation Trust, Cardiff University, the University of Stirling, and the London School of Hygiene & Tropical Medicine (“we”, “us” and “our”) are committed to protecting your personal data.

The notice is addressed to individuals who are taking part in the research study entitled ‘The Supporting MumS (SMS) study’ (“you” and “your”).

This Privacy Notice tells you why we need to collect personal information about you, what we will do with it, and how we will look after it. It also tells you about your legal rights in relation to your Personal Data.

If you have any questions about this privacy notice, please contact us. Contact details are provided below.

**WHO WE ARE**

1. We are a group of expert researchers from the organisations listed above. The study is led by Queen’s University Belfast and is funded by National Institute of Health Research and the Public Health Agency Northern Ireland.
2. All our research is underpinned by policies and procedures that ensure we comply with regulations and legislation that govern the conduct of research. This includes data protection legislation: the UK General Data Protection Regulation (GDPR) and the Data Protection Act 2018 (DPA). In the case of health and social care research, which should serve the public interest, where we have to demonstrate that our research serves the interests of society as a whole.

**HOW YOUR PERSONAL DATA IS COLLECTED**

3. **Information you provide:** When you consent to take part in the SMS study we will ask for information about you, such as your name, address, NHS number, date of birth, telephone number, email address and date of birth of your youngest child. This is known as your “Personal Data”. We may also ask you for some special categories of information (for example information about your physical and mental health). This is known as your “Sensitive Personal Data” and has additional protections.
4. **Data from other sources:** We will also collect information about you from other sources and this also forms part of “Personal Data” and “Sensitive Personal Data”. This includes information from:

- the system we are using to send and receive text messages for the study, for example we will automatically collect the replies you send to the text messages. This system is owned and operated by the London School of Hygiene & Tropical Medicine.
- routine health-related data (if you have consented to this option) such as NHS records, GP and dental records, maternity and health visiting records and disease registers. To obtain your routine data from these systems, we will share some personally- identifiable details such as name, date of birth, address and NHS number to each of these organisations and they will send us your data back. This will all be done using secure data transfer systems that have been set up carefully to keep your information safe and complies with regulations and legislation governing the conduct of research.

**HOW WE USE YOUR PERSONAL DATA**

5. We use your Personal Data and Sensitive Personal Data in the following ways:

- to contact you to invite you to take part in the study and to discuss research-related queries. If you do not consent to take part in the study, your personal data, such as contact details (e.g. name, address, phone number), will be confidentially destroyed and will not be retained by us.
- after consent to take part is provided, personal data is used to contact you to arrange and conduct study visits, to request information or to send you relevant study documents or equipment and to inform you about any relevant study developments.
- access to, and sharing of, special category and sensitive personal data is controlled very carefully. Sensitive personal data is collected and pseudonymised – this means each person who takes part in this study will be given a unique study number and this unique study number is used on all the study paperwork and in study databases. Only authorised members of the SMS research team will be able to link your study number to your name, contact details and NHS number.

6. In order to protect your rights and freedoms when using your personal information for research and to process special category information we have special safeguards in place to protect your information, and all information is kept in line with our policies and regulatory requirements.
7. In addition to the above safeguards, data protection legislation requires us to meet the following standards when we conduct research with your personal information:

- the research will not cause damage or distress to someone (e.g., physical harm, financial loss or psychological pain).
- the research is not carried out in order to do or decide something in relation to an individual person, unless the processing is for medical research approved by a research ethics committee.
- the Data Controller has technical and organisational safeguards in place (e.g. appropriate staff training and security measures).
- if processing a special category of data, this must be subject to a further public interest test to make sure this particularly sensitive information is required to meet the research objectives.

**LEGAL BASIS FOR COLLECTING AND USING YOUR PERSONAL DATA**

8. We will only use your Personal Data if we have valid reasons for doing so. These reasons are known as our “legal basis for processing”.
9. In the context of research, the lawful basis upon which we will process your personal information is usually where “Processing is necessary for the performance of a task carried out in the public interest or in the exercise of official authority vested in the controller” (Article 6 of GDPR).
10. We also collect and use more sensitive personal information (Special Category data) where “the processing is necessary for archiving purposes in the public interest, scientific or historical research purposes or statistical purposes… which shall be proportionate to the aim pursued, respect the essence of the right to data protection and provide for suitable and specific measures to safeguard the fundamental rights and the interests of the data subject”. (Article 9 of GDPR).

**WHO WE SHARE YOUR DATA WITH**

11. In line with our Data Protection Policy and Procedures we can share your information, including Personal Data and Sensitive Personal Data, with the following parties for the following research purposes:

- London School of Hygiene and Tropical Medicine (LSHTM) text message system – to allow you to receive and reply to the text messages (the LSHTM is a study collaborator)
- Designated research personnel for the purposes of achieving the research outcomes
- The Northern Ireland Clinical Trials Unit (NICTU) for the purposes of entering your completed questionnaires onto a study database (NICTU is a study collaborator)
- Qualtrics research software supplier for the purposes of the online completion of the study questionnaires (privacy notice can be viewed here - <https://www.qualtrics.com/support/survey-platform/getting-started/data-protection-privacy/>).
- Transcription services for the purpose of transcribing recorded interviews (if consent provided for this optional element)
- Organisations in-charge of the collection and administration of routine data for the purposes of achieving the research outcomes (if consent provided for this optional element)

12. In all cases, information shared will be on a need to know basis, not excessive and with all appropriate safeguards in place to ensure the security of your information.
13. When we use third parties (known as data processors) to carry out a task on our behalf, such as a transcription service as indicated above, we have contractual terms, policies and procedures to ensure confidentiality is respected.

**DATA PROCESSING OUTSIDE EUROPE**

14. We will not transfer your Personal Data and Sensitive Personal Data outside of the United Kingdom and European Economic Area.

**HOW LONG YOUR INFORMATION WILL BE KEPT?**

15. Information where you can be identified will be kept for a minimum amount of time and in accordance with research objectives. Researchers will de-identify information, i.e. anonymise or pseudonymise, as soon as possible as described in point 5.
16. We will keep your Personal Data and Sensitive Personal Data for up to 10 years from when the research has been completed to allow for full and final publication of the research results. If you have provided consent (this is optional), we will retain your record of consent and personal data (name, telephone number, email address, address) for the purpose of conducting longer-term follow-up for the SMS study or to inform you about future studies related to health and lifestyle that we may be conducting. If you have consented, we will retain your NHS number for data linkage purposes. You can decline to receive communications at any stage. We will only keep your information if we need it for one of the reasons described above.
17. We place great importance on the security of the Personal Data that we hold, including the use of physical, technological and organisational measures to ensure your information is protected from unauthorised access and against unlawful processing, accidental loss, alteration, disclosure, destruction and damage.

**YOUR RIGHTS**

18. The DPA provides you with a number of legal rights in relation to your Personal Data, including the right:

- to request access to your Personal Data;
- to request correction of your Personal Data that is wrong or incomplete;
- to request erasure or the restriction of processing of your Personal Data;
- to request the transfer of your Personal Data in a structured; commonly used machine-readable format;
- not to be subject to automated decision making; and
- to withdraw your consent.

19. If you wish to exercise any of the rights set out above, or require further information about any of the rights, please contact us.
20. There may also be times where we cannot stop using your Personal Data when you ask us to, but we will tell you about this if you make a request.

**CONTACTING US**

21. If you have any questions or comments about this privacy notice, please contact the SMS team:

Dr Dunla Gallagher

SMS Trial Manager

Centre for Public Health

Institute Clinical Science A

School of Medicine, Dentistry & Biomedical Sciences

Queen’s University Belfast

Professor Michelle McKinley

SMS Chief Investigator

Centre for Public Health

Institute Clinical Science A

School of Medicine, Dentistry & Biomedical Sciences

Queen’s University Belfast

Alternatively, you can contact:

Data Protection Officer

Information Compliance Unit
Lanyon South
Queen’s University Belfast
University Road
BT7 1NN

**COMPLAINTS**

22. You have the right to complain about how we treat your Personal Data and Sensitive Personal Data to the Information Commissioner’s Office (ICO). The ICO can be contacted at:

Information Commissioner's Office
Wycliffe House
Water Lane
Wilmslow
Cheshire
SK9 5AF

**CHANGES TO THIS NOTICE**

23. We may update this Privacy Notice from time to time. We will notify you of the changes where we are required by law to do so.

c) **Study Consent Form**

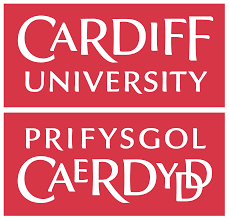

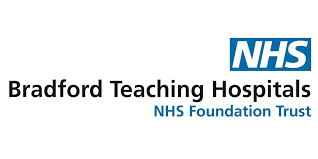

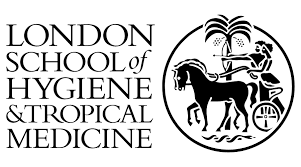

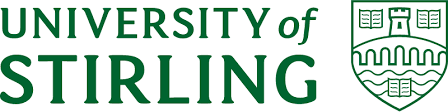

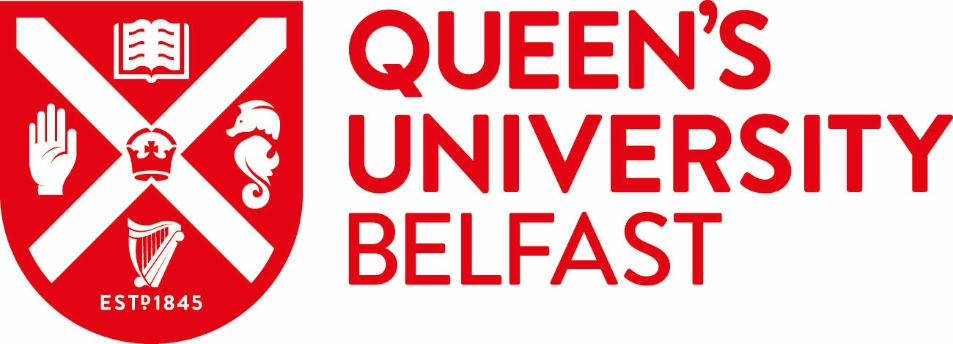

**CONSENT FORM**

**The Supporting MumS (SMS) study**

**Screening Number: __ __ __ __ __	Participant ID number: __ __ __ __ __**

|  |  | Please **initial** box |
| --- | --- | --- |
| 1. | I confirm that I have read and understood the Participant Information Sheet dated 05/04/2022 (Version 4.0) and privacy notice dated 30/11/2021 (Version 1.0) for the above study and have been given copies to keep. | 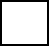 |
| 2. | I have had the opportunity to consider the information, ask questions and have had these answered to my satisfaction. | 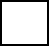 |
| 3. | I understand that my participation is voluntary and that I am free to withdraw at any time, without giving any reason and without my medical care or legal rights being affected. | 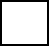 |
| 4. | I understand that information collected about me for the study (including personally identifiable information) may be looked at by responsible individuals in the study team and regulatory authorities supervising the study. | 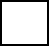 |
| 5. | I understand that all information collected about me during the course of the research will be kept strictly confidential and processed in compliance with data protection legislation. | 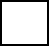 |
| 6. | I understand that some of the data collected about me will be processed by third parties (known as data processors), such as software and transcription companies, and that contractual terms, policies and procedures will be put in place to ensure confidentiality is respected. | 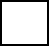 |
| 7. | I give permission for my GP to be informed of any concerning medical issues detected during study visits. | 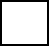 |
| 8. | I am aware of the potential risks and benefits of this research study, as described in the Participant Information Sheet and discussed with the researcher. | 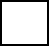 |
| 9. | I understand that I will not be identifiable in any published report using data from this study. | 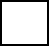 |
| 10. | I agree to my contact details being kept so I can be informed of the study findings. | 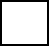 |
| **11.** | **I consent to take part in the above study.** | 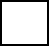 |
|  | **OPTIONAL ELEMENTS (please initial box if you consent)** |  |
| 12. | I am willing to be contacted to take part in two short (20-30 minute) telephone interviews during the study to provide feedback to the research team. | 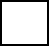 |
| 13. | I am willing to be contacted in the future about a follow-up to this study. | 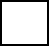 |
| 14. | I consent to be contacted by the University about future research studies related to diet or lifestyle and health for which I may be eligible. | 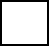 |
| 15. | I consent to my personal health information, including my NHS number and date of birth, being released to organisations that hold routine health-related data so they can locate information about me that is held in their database. | 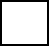 |
| 16. | I consent to organisations holding my routine health-related data performing data linkage and releasing the information about me from its databases to the SMS Study research team. | 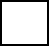 |

| Name of Participant (Block capitals) | Signature | Date |
| --- | --- | --- |

**To be completed by the Principal Investigator or nominee**

**I, the undersigned, have taken the time to fully explain to the above patient the nature and purpose of the study in a way that they could understand. I have explained the risks involved as well as the possible benefits. I have invited them to ask questions on any aspect of the study that concerned them.**

| Name of Researcher (Block capitals) | Signature | Date |
| --- | --- | --- |

| Researchers’ Name, Address, Email address, Telephone (site specific) | Dr Dunla Gallagher  SMS Trial Manager  Centre for Public Health, Queen's University Belfast School of Medicine, Dentistry & Biomedical Sciences  Institute of Clinical Sciences (Block B)  Grosvenor Road, Belfast, N.Ireland, BT12 6BA  Tel: 07341 888415  |
| --- | --- |

**Supplemental appendix 5- World Health Organization Trial Registration Data Set**

| **Data Category** | **Information** |
| --- | --- |
| **Primary Registry and Trial Identifying Number** | ISRCTN Registry; ISRCTN16299220 |
| **Date of Registration in Primary Registry** | 10/02/2022 |
| **Secondary Identifying Numbers** | FUNDERS Number: NIHR131509  Sponsor Number: B21/24  IRAS Number: 305557  REC Reference: 22/WS/0003 |
| **Source(s) of Monetary or Material Support** | National Institute for Health and Care Research (NIHR) Public Health Research (PHR). Intervention costs provided by the Public Health Agency (Northern Ireland). |
| **Primary Sponsor** | Queen’s University Belfast |
| **Secondary Sponsor(s)** | None |
| **Contact for Public Queries** | Dr Dunla Gallagher  SMS Trial Manager   07341 888415  Centre for Public Health  Queen’s University Belfast  Institute of Clinical Sciences B  Grosvenor Road  Belfast  BT12 6BJ |
| Contact for Scientific Queries | Professor Michelle McKinley  Chief Investigator   028 9097 8936  Centre for Public Health  Queen’s University Belfast  Institute of Clinical Sciences A  Grosvenor Road  Belfast  BT12 6BA |
| **Public Title** | The Supporting MumS (SMS) study: can we use an automated text message intervention for weight management in women after childbirth? |
| **Scientific Title** | Effectiveness and cost effectiveness of an automated text message intervention for weight management in postpartum women with overweight or obesity: the Supporting MumS (SMS) Randomised Controlled Trial. |
| **Countries of Recruitment** | United Kingdom (Northern Ireland, Scotland, England and Wales). |
| **Health Condition(s) or Problem(s) Studied** | Overweight and obesity in postpartum women. |
| **Intervention(s)** | Supporting MumS intervention group- will receive automated text messages about weight loss and maintenance of weight loss for 12 months. The text messages will focus on diet and physical activity with embedded behaviour change techniques known to be positively associated with weight management.  Active control group- will receive automated text messages about child health and development for 12 months. |
| **Key Inclusion and Exclusion Criteria** | Inclusion criteria   - Women (as per NICE Postnatal care guideline NG194^1^, the term 'woman' is taken to include people who do not identify as women but who are pregnant or have given birth) - Aged > 18 years old - BMI ≥25 kg/m^2^ - Have had a baby within the last two years   Exclusion criteria   - Baby less than 6 weeks old - No access to a mobile phone to receive personal messages - Insufficient English to understand short written messages - Currently pregnant - Recent or planned bariatric surgery - Eating disorder - On a specialist diet and receiving dietetic care - Taking part in another weight management research study currently, or in the last 3 months |
| **Study Type** | Two-arm parallel groups randomised controlled trial to test the effectiveness and cost-effectiveness of a 12-month text-messaging behavioural weight management intervention in supporting weight loss for women with overweight and obesity in the postpartum period, compared with an active control group receiving text messages related to child health and development for 12 months.  Women will be recruited from all four countries in the UK using community-based recruitment as well as signposting via routine contact with health professionals.  Written informed consent will be obtained, baseline data collected and then participants will be randomised. Participants will be block randomised and randomisation will be stratified by site. The randomisation sequence will be developed in STATA by a statistician who is independent of the study team. The randomisation will be implemented via the London School of Hygiene and Tropical Medicine secure remote web-based system which will link directly with the text message database and will deliver the intervention or active control content according to the random allocation sequence.  Participants will become aware of their group allocation when they start to receive the messages. Researchers who are recruiting women to the study and collecting outcome data are blinded to group allocation. |
| **Date of First Enrolment** | Actual date of enrolment of the first participant: 20/05/2022 |
| **Sample Size** | Target sample size: 888  Final number of participants that the trial enrolled: 892 |
| **Recruitment Status** | Complete: participants are no longer being recruited or enrolled. |
| **Primary Outcome** | Weight change from baseline to 12 months (kg), measured using calibrated scales. |
| **Secondary Outcome** | 1. Waist circumference measured by a flexible measuring tape at baseline, 6, 12 and 24 months, and mean BMI (kg/m^2^) and the proportions of women gaining a substantial amount of weight (>5kg) at 12 and 24 months.  2. Health behaviours are measured by: Fat and Fibre Barometer; self-report questionnaire on sugar and alcohol intake; International Physical Activity Questionnaire (IPAQ) - Short form; and self-report questionnaire on infant feeding practices at baseline, 6, 12 and 24 months.  3. Study acceptability is measured by: recruitment; retention; engagement with the two-way text messages over 12 months; self-reported questionnaire on participant satisfaction with SMS messages (at 6 and 12 months) and rating of intervention (at 12 months); and qualitative interviews (at 6 and 12 months).  4. Economic evaluation outcomes are measured by: self-report questionnaire on health service resources use, medication usage and lifestyle-related costs; the EuroQol 5-dimension (EQ-5D) quality of life questionnaire; and the ICEpop Capability Measure for Adults (ICECAP-A) at baseline, 6, 12 and 24 months.  5. Moderators of intervention effect are measured by: Edinburgh Postnatal Depression Scale (EPDS); Generalised Anxiety Disorder (GAD-7); Pittsburgh Sleep Quality Index; and self-report questionnaire on confidence and desire for weight loss and maintenance at baseline, 6, 12 and 24 months.  6. Mediators of intervention effect are measured by: Health Action Process Approach (HAPA) for diet and exercise; Self-Report Behavioural Automaticity Index for diet and exercise; Self-regulation of eating behaviour questionnaire; self-report questionnaire on monitoring and goal setting for diet and exercise; Motivation for weight loss scale; Social support for eating & exercise questionnaire; and Rosenberg Self-Esteem Scale at baseline, 6, 12 and 24 months. |
| **Ethics Review** | Approved 24/02/2022 (Ref: 22/WS/0003) by West of Scotland REC 4 (West of Scotland Research Ethics Service, Ward 11, Dykebar Hospital, Grahamston Road, Paisley, PA2 7DE, United Kingdom; +44 (0)141 3140213;). |
| **Completion date** | 30/04/2025 |
| **Summary results** | Intention to publish date: 30/04/2026. |
| **IPD Sharing Statement** | Available on request. Formal requests to be made in writing to the CI (Prof M McKinley;). |
